# Individualised and Time-Dependent Prediction of Recurrence After Colorectal Cancer Liver Metastases Resection

**DOI:** 10.64898/2026.11.04.26367191

**Authors:** Annika Viljamaa, Ruth Baumgartner, Rickard Strandberg, Johanna Sterner, Sarkis Drejian, Davit Aghayan, Carlos Fernández Moro, Ernesto Sparrelid, Pia Osterlund, Bjørn Edwin, Åsmund A. Fretland, Natalie Geyer, Jennie Engstrand, Marco Gerling

**Author notes:** equal contributions, corresponding authors (Jennie Engstrand, Marco Gerling). **Correspondence to:** Jennie Engstrand, MD, PhD, Marco Gerling, MD, PhD, CLINTEC, Karolinska Institutet, C1:77 14186 Huddinge, Sweden.

## Abstract

**Introduction:** Treatment strategies for colorectal cancer liver metastases (CRLM) are increasingly complex, driving interest in advanced prognostic tools. Externally validated benchmarks based on established clinical risk scores are needed to contextualise emerging models. However, existing scores remain difficult to apply and compare, because they provide categorical risk strata rather than individualised probabilities, and because model parameters are rarely published. Here, we validate and recalibrate seven established risk scores to derive individualised recurrence probabilities for future score benchmarking.

**Method:** Patients undergoing index CRLM resection from two tertiary centres were included: Karolinska University Hospital (retrospective KaroLiver cohort [2013–2021, follow-up to 2025, n=709]) and Oslo University Hospital (OSLO-COMET trial cohort [2012–2016, follow-up to 2022, n=198]). Performance of the GAME, m-CS, Fong, Nordlinger, Nagashima, Konopke and Helsinki scores was assessed using recurrence-free survival as the primary outcome.

**Results:** Among 709 patients in KaroLiver (median age 67 years [IQR 59–74]; 434 men [61%]), 422 (60%) had synchronous metastases and 444 (63%) received neoadjuvant chemotherapy. All scores significantly stratified recurrence risk (log-rank P<0.001; time-dependent AUC 0.63–0.74 at 3 years), albeit with performance differences across time points. Recalibrated models showed good agreement between predicted and observed recurrence and were externally validated in OSLO-COMET with broadly comparable net benefit. Models were implemented in the online tool PLATO (Personalised Liver metastases risk AssessmenT in Oncology).

**Conclusion:** Established CRLM risk scores can be translated into individualised probabilistic recurrence estimates at clinically relevant time points, supporting patient communication, risk-adapted surveillance and transparent benchmarking of next-generation prognostic models.

## Introduction

Despite curative-intent surgery, recurrence after resection of colorectal cancer liver metastases (CRLM) remains a major clinical challenge, affecting up to 70% of patients^1–5^. Management of CRLM has become increasingly complex as patient selection, surgical approaches, and systemic therapies have evolved^6^. Surgical strategies have expanded toward parenchymal-sparing techniques and thermal ablation^7,8^, while liver transplantation is now considered for selected patients^9,10^. Concurrently, systemic therapies are becoming increasingly effective^11^. Although these advances are currently limited to subgroups of patients, they underscore a general shift in the management of CRLM: while technical resectability remains the central criterion, clinical decision-making increasingly requires integrating tumour biology and patient-specific factors into a multidisciplinary framework, ideally informed by individualised estimates of recurrence risk.

Various prognostic scores have been developed to predict recurrence and survival after CRLM resection^12–16^, and many have undergone external validation in contemporary cohorts^17–21^. Nevertheless, these scores are rarely integrated into routine practice due to specific hurdles to their implementation: most existing models express risks as ordinal scores or categorical strata rather than time-specific probabilities, limiting their interpretability. Moreover, performance evaluation is often restricted to measures of discrimination, without assessment of calibration or performance at clinically meaningful time points^22^. Even when models have been externally validated, the underlying regression coefficients and model intercepts are rarely reported, precluding direct recalibration and adaptation to new populations. Hence, novel prognostic tools based on emerging methods such as circulating tumour DNA^23,24^ or advanced radiological image analyses^25–27^ can only be indirectly compared with existing clinical scores, and efforts to compare and integrate established models with novel approaches have been limited.

Here, we evaluate seven clinical risk scores in a comprehensive framework that included discrimination, calibration, and net benefit analyses at multiple clinically relevant time points. We translate them into individualised, percentage-based recurrence probabilities, and publish all model parameters and source code through an open-source online calculator to enable future adaptation and benchmarking.

## Methods

### Study Design and Setting

For score validation and model recalibration, this retrospective observational study was based on the KaroLiver cohort, which included consecutive patients 18 years or older who underwent curative-intent liver intervention at the tertiary centre, Karolinska University Hospital, Stockholm, Sweden, from 2013 to 2021 (follow-up to 2025). Validation was performed in the OSLO-COMET randomised controlled trial^28^, which included patients from 2012 to 2016 (follow-up to 2022) at the tertiary centre, Oslo University Hospital, Oslo, Norway^28^. Only index resections were included (**Fig. S1**).

The study was conducted in accordance with the Declaration of Helsinki and reported in accordance with the Strengthening the Reporting of Observational Studies in Epidemiology (STROBE) guideline^29^. KaroLiver was approved by the Swedish National Ethical Review Authority (EPM 2019-01571, amendments 2020-00428, 2021-06863-02, 2022-06588-02, and 2025-05423-02); informed consent was waived. OSLO-COMET was approved by the Regional Ethics Committee of Southeastern Norway (REK Sør-Øst B 2011/1285); all participants provided written informed consent.

The study was not preregistered, and the study protocol was not published prior to the study. Patients or patient advisory bodies were not involved in the study.

### Participants

KaroLiver included patients with histologically confirmed colorectal carcinoma and radiologically confirmed liver metastases. Patients were reviewed by a liver-specific multidisciplinary team (MDT); in the event of disease recurrence, MDT re-evaluation was performed, with curative-intent retreatment pursued when technically feasible and oncologically appropriate. Patients treated with ablation only were not included. Patients with extrahepatic disease were included when amenable to complete curative-intent treatment. To meet the assumption of full outcome observation for all subjects in generalised linear models, 28 patients who were lost to follow-up at 3 years were excluded from model recalibration (**Fig. S1a**).

Inclusion criteria for OSLO-COMET have been previously described (**Fig. S1b)**^3,28^; it enrolled patients amenable to parenchyma-sparing resection, comparing open with laparoscopic techniques.

For both cohorts, chemotherapy was administered according to the local guidelines in effect at the time of each patient’s treatment, principally aligned with the European Society for Medical Oncology (ESMO) Guidelines.

Postoperative surveillance consisted of thoracic and abdominal contrast-enhanced computed tomography (CT) and tumour marker measurements at the treating physician’s discretion. Baseline recommendations differed: in KaroLiver, every 3 to 6 months; in OSLO-COMET, every 4 months until 2 years after surgery, then every 6 months.

### Data collection

For KaroLiver, data were retrospectively extracted from electronic patient records. Variables included patient age and sex, primary metastases tumour characteristics. Mutational and microsatellite instability/mismatch repair testing were performed on a per-demand basis in clinical routine. Data for OSLO-COMET were collected as previously described^28^.

### Variable and Outcome Definitions

In KaroLiver, disease-free interval (DFI) was defined as the time between diagnosis of CRC and CRLM, and synchronous disease as DFI <1 month. In OSLO-COMET, DFI was recorded to indicate whether CRLM was diagnosed within 12 months of CRC diagnosis. Recurrence-free survival (RFS) was defined as the time from CRLM resection to first confirmed recurrence, including local or distant recurrences documented radiologically, endoscopically, or clinically. Patients alive and without confirmed recurrence were censored on the date of last imaging in KaroLiver, or last documented follow-up in OSLO-COMET.

Established clinical risk scores were calculated according to the original score definitions. These included extensively validated and cited scores^20^ (Fong, Konopke, Nordlinger, Nagashima)^12,13,15,30^, and recent scores incorporating tumour-biological variables such as histology (Helsinki)^14^ and/or genetic variant background (Genetic And Morphological Evaluation [GAME] score, modified clinical score [m-CS])^31,32^. Of note, RFS and OS correlate only moderately after CRLM resection, necessitating deliberate selection of the primary outcome^33^. Given the heterogeneous disease trajectories in this population and the clinical relevance of recurrence timing for, e.g., surveillance interval planning, RFS was chosen as the primary outcome.

### Statistical Analysis

Statistical analyses were performed in R v4.4.1^34^. Statistical tests were 2-tailed, and *P*<0.05 was considered significant. Missing data were imputed using multiple imputation by chained equations^35^. The scores’ performances were evaluated by discrimination, calibration, and clinical utility analyses at 6 months, 1, 2, and 3 years in accordance with the Transparent Reporting of a Multivariable Prediction Model for Individual Prognosis or Diagnosis (TRIPOD) checklist for prediction model verification (**Table S1**)^36^.

### Recalibration of Scoring Models

Recalibration was performed by fitting logistic regression models that link the scoring factors to recurrence status, translating categorical score outcomes onto a probabilistic scale. Internal validation of the recalibrated models was performed using bootstrapping in KaroLiver; models were externally validated in OSLO-COMET.

### Online Prediction Tool

PLATO was developed as a Shiny v1.11 application^37^ in R, based on logistic regression model coefficients from model recalibration.

Full details of statistical analyses, model development, and technical details of PLATO are provided in the **Supplementary Material.** The code and the Shiny framework are available at https://github.com/vilja-a/plato-crlm-risk-prediction/. Score parameters and model coefficients are publicly reported.

## Results

### Validation of seven risk scores to predict recurrence after CRLM resection

We followed a stepwise process: using a real-world consecutive cohort, we (1) validated established clinical risk scores, (2) recalibrated them to provide individualised risk percentages, and (3) developed a web-based calculator that (4) was validated in an external cohort (**Fig. 1a**).

**Fig. 1.**
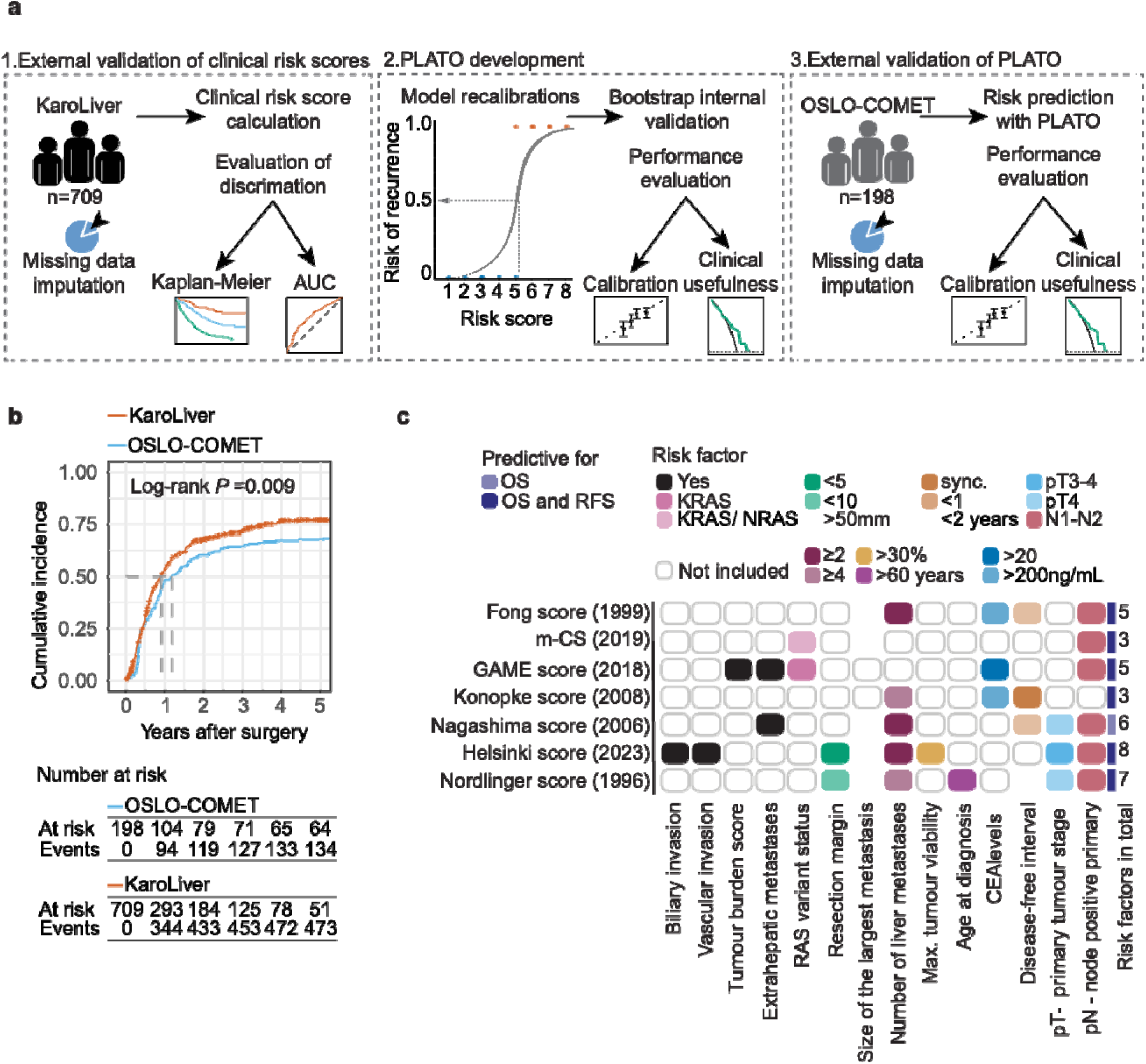
Study Design, Analysis Cohorts, and Description of Clinical Risk Scores. **a** Schematic of the study workflow. **(1)** KaroLiver, a real-world surgical cohort, was used to evaluate the discrimination of seven established clinical risk scores. **(2)** Models were recalibrated into a probabilistic scale using logistic regression, and PLATO was created; recalibrated model performance was evaluated after internal validation. **(3)** Final validation was performed in the independent OSLO-COMET cohort. **b** Cumulative incidence of recurrence. Patients at risk, *P* value, statistical test, and stratum definition are displayed in the plots. **c** Description of the clinical risk scores and scoring variables. AUC, area under the curve; CEA, carcinoembryonic antigen; CRLM, colorectal cancer liver metastasis; OS, overall survival; PLATO, personalised liver metastases risk assessment in oncology; sync., synchronous; RFS, recurrence-free survival.

#### Study Cohorts

In KaroLiver, 709 patients (median age 67 years [IQR, 59-74 years]; 434 males [61%]) underwent their index CRLM resection. A total of 422 patients (60%) had synchronous metastases, and 444 (63%) received neoadjuvant chemotherapy. Major hepatectomy was performed in 313 patients (44%). Information on mutational status was available for 314 patients (44%). Despite differences in patient selection, recurrence timing was broadly comparable between cohorts, while the recurrence rate was lower in OSLO-COMET (median RFS 14.3 [95%CI 11.1-20.0] months vs 10.9 [95%CI 9.2-12.4] months; recurrence rates 6.0% vs 12.3%; *P*=0.009; **Fig. 1b**). OSLO-COMET patients had fewer liver metastases (median 1 vs 2; *P*<0.001) and received neoadjuvant chemotherapy less frequently (42.4% vs 63.2%; *P*<0.001). Baseline characteristics are given in **Table 1**.

**Table 1.** Clinical characteristics of the KaroLiver and OSLO-COMET cohorts

| **Clinical Parameter** | **KaroLiver (N=709)** | **OSLO-COMET (N=198)** | ***P* Value** |
|---|---|---|---|
| **Age at liver resection, median (IQR), years** | 67 (59, 74) | 67 (60,73) | 0.99 |
| **Male sex, No. (%)** | 434 (61.2) | 119 (60) | 0.84 |
| **Mutational status, No. (%)** |  |  |  |
| *KRAS, NRAS* wild type | 156 (49.8) | 24 (50.0) | 0.19 |
| *KRAS* mutation | 146 (46.7) | 21 (43.7) |  |
| *NRAS* mutation | 10 (3.2) | 3 (6.3) |  |
| *KRAS* and *NRAS* mutations | 1 (0.3) | 0 |  |
| Missing *NRAS* or *KRAS* status | 396 | 150 |  |
| *BRAF<sup>V600E</sup> wild type* | 269 (91.8) | 46 (95.8) | 0.49 |
| *BRAF<sup>V600E</sup>* mutation | 24 (8.2) | 2 (4.2) |  |
| Missing *BRAF<sup>V600E</sup>* status | 416 | 150 |  |
| **MSI/dMMR** | 10 (6.8) | - |  |
| *Missing* | 561 | - |  |
| **Primary colorectal cancer** |  |  |  |
| **Primary tumour location, No. (%)** |  |  |  |
| Right-sided | 181 (25.8) | 35 (18.5) | 0.048 |
| Left-sided, including rectum | 521 (74.2) | 154 (81.5) |  |
| *Missing* | 7 | 9 |  |
| **pT stage primary tumour, No. (%)** |  |  |  |
| T0* | 9 (1.4) | 0 | 0.19 |
| T1-T2 | 88 (14) | 14 (8) |  |
| T3 | 317 (50) | 125 (70) |  |
| T4 | 223 (35) | 40 (22) |  |
| *Missing* | 72 | 19 |  |
| **pN stage primary tumour, No. (%)** |  |  |  |
| N0 | 200 (31) | 55 (31) | 0.99 |
| N1-N2 | 436 (69) | 122 (69) |  |
| *Missing* | 73 | 21 |  |
| **Liver metastases** |  |  |  |
| **Time of detection** |  |  |  |
| Synchronous No. (%) | 422 (59.5) | 127 (64.1) | 0.27 |
| *Missing* | 0 | 0 |  |
| **Number of liver metastases, No. (%)** |  |  |  |
| 1 | 266 (37.5) | 113 (57.1) | <0.001 |
| 2-5 | 301 (42.5) | 82 (41.4) |  |
| 6-10 | 91 (12.8) | 3 (1.5) |  |
| 11 or more | 51 (7.2) | 0 |  |
| *Missing* | 0 | 0 |  |
| **Size of the largest liver metastasis, median (IQR), mm** | 25 (15, 40) | 24 (15,35) | 0.13 |
| *Missing* | 4 | 1 |  |
| **Simultaneous lung metastases, No. (%)** | 83 (11.7) | - |  |
| *Missing* | 1 | - |  |
| **Extrahepatic, non-pulmonary metastases, No. (%)** | 42 (5.9) | - |  |
| *Missing* | 0 | - |  |
| **Neoadjuvant chemotherapy, No. (%)** | 444 (63.2) | 84 (42.4) | <0.001 |
| *Missing* | 7 | - |  |
| **CEA at liver resection, median, (IQR), µg/L** | 6 (3, 21) | 4 (2, 9) | <0.001 |
| *Missing* | 402 | 5 |  |
| **Resection margin, median (IQR), mm** | 1 (0, 4) | 3 (1, 7) | <0.001 |
| *Missing* | 51 | - |  |
| **Vascular invasion, No. (%)** | 316 (51.3) | - |  |
| *Missing* | 93 | - |  |
| **Biliary invasion, No. (%)** | 148 (24.1) | - |  |
| *Missing* | 96 | - |  |
| **Tumour maximum viability, median (IQR), %** | 50 (25, 70) | - |  |
| *Missing* | 284 | - |  |

\*T0: after neoadjuvant treatment (ypT)

Imputed and original data distributions were statistically similar; the lowest *P* value was observed for *NRAS* status (set #2; 4.8% vs 7.6%; *P*=0.14 KaroLiver, and set #3 6.3% vs 14.7%; *P*=0.136 OSLO-COMET; **Fig. S2 & S3**).

#### Recurrence and Survival in the KaroLiver cohort

After a median follow-up of 44.9 months (95% CI, 41.1-49.5 months), recurrence occurred in 467 of 709 patients (65.9%), most commonly in the liver (191 of 467 [40.9%]) and lung (98 of 467 [20.9%]). The median time to recurrence was 7.1 months (95% CI, 6.5-7.6 months). The 1-year and 3-year RFS rates were 46.5% (95% CI, 42.8%-50.6%) and 28.3% (95% CI, 24.9%-32.1%), respectively. Median OS was 51 months (95% CI, 46.1-59.6 months, compared with the median RFS of 10.9 months [95%CI 9.2-12.4]). The 3-year and 5-year OS rates were 61.3% (95% CI, 57.8%-65.1%) and 45.6% (95% CI, 41.8%-49.8%), respectively.

#### Score Validation - Discrimination

Seven established clinical risk scores predictive of RFS and/or OS were validated (**Fig. 1C**). Time-dependent AUC analyses revealed improved performance at later time points after surgery. At 6 months, the Nordlinger score nominally showed the highest AUC (0.72); at 1 year, the Helsinki score (0.69); and from 1.5 years onwards, the GAME score (0.72-0.76; **Fig. 2a**).

**Fig. 2.**
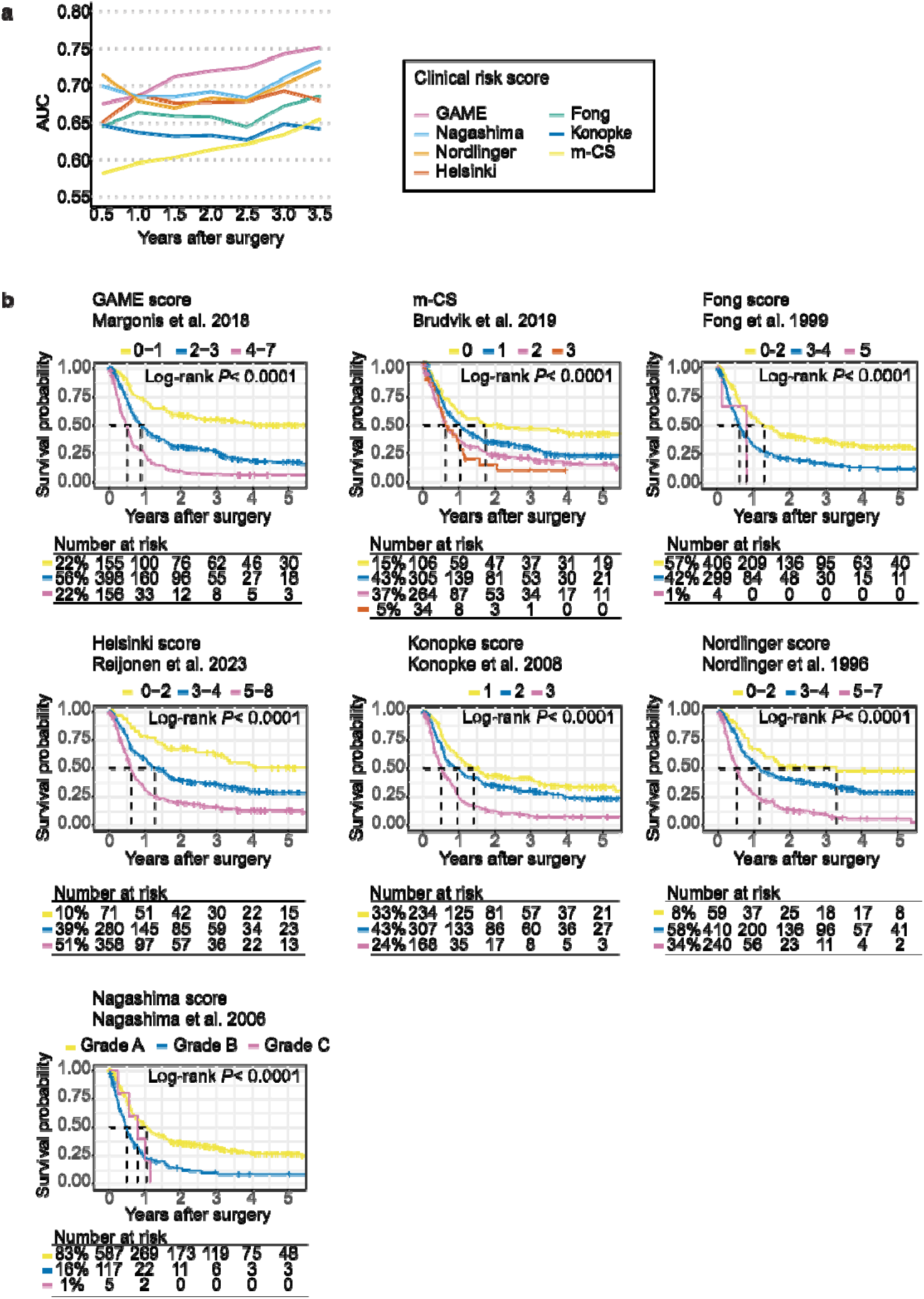
Discrimination of Recurrence-Free Survival by Clinical Risk Scores in KaroLiver. **a** Time-dependent area under the curve (AUC) for recurrence-free survival (RFS) by clinical risk score. **b** Kaplan-Meier estimates of RFS stratified by risk scores, based on original strata. Patients at risk, *P* values, statistical tests, and stratum definitions are displayed in the plots. CRLM, colorectal liver metastasis; GAME, genetic and morphological evaluation; m-CS, modified clinical risk score.

All scores significantly stratified RFS (*P*<0.001; **Fig. 2b**), albeit with markedly different distributions across low- and high-risk categories. In low-risk groups, long median RFS was reached among Nordlinger, GAME and Helsinki scores: using the originally published strata, the Nordlinger score classified 59 of 709 patients (8.3%) as low risk with median RFS of 39.6 months, GAME 155 of 709 patients (21.9%), with median RFS of 82.5 months, and Helsinki 71 of 709 patients (10.0%), with median RFS not reached.

Konopke and Nordlinger scores classified 168 of 709 patients (23.7%) and 240 of 709 patients (33.8%), respectively, as high risk, with a median RFS of 6.1 months for both. Among more recent scores, the Helsinki, m-CS, and GAME scores classified 156 (22.0%), 254 (35.8%), and 358 of 709 patients (50.5%) as high risk, with median RFS of 6.0, 7.2, and 7.6 months, respectively. The highest-risk classes of the Nagashima, Fong, and m-CS scores showed limited discriminative utility using their pre-defined strata, classifying only 4 (0.6%), 5 (0.7%), and 34 of 709 patients (4.8%), respectively.

Intermediate-risk classification was the most consistent across the scores, including 280 (39.5%) to 410 (57.8%) of 709 patients, with median RFS ranging from 10.6 to 15.4 months. The Nagashima score was an exception, classifying fewer patients (117 of 709 [16.5%]) as intermediate risk with a notably shorter median RFS of 6.0 months.

Despite a known moderate association between RFS and OS after CRLM resection^33^, which was confirmed in KaroLiver (Kendall’s t 0.52, 95%CI [0.48-0.57], *P*<0.001), all scores also significantly stratified OS (log-rank *P*<0.001; **Fig. S4**). Moreover, in multivariable analyses, the scoring factors were independently associated with RFS and OS (global *P* values < 0.001; **Fig. S5-S8**).

### Calibration and Score Performance at Distinct, Clinically Meaningful Time Points

To evaluate score performance with calibration and clinical utility assessments at relevant time points, we recalibrated the scoring models to a probabilistic scale (**Fig. 1a, box 2**). Inspection of calibration plots confirmed successful recalibration: mean observed event rates closely followed a 45-degree line, indicating overall agreement between predicted and observed risks (**Fig. S9**).

Most scores demonstrated good agreement between predicted and observed recurrence probabilities, with observed-to-expected ratios close to unity across scores and time points in KaroLiver after internal validation, and in the OSLO-COMET cohort (**Fig. 3, Fig. S10, Table S2**). At 6 months, Fong, Nagashima, and Nordlinger scores overestimated recurrence risk at higher predicted probabilities (>50%) in KaroLiver. Although agreement between observed and predicted risks was generally good at 1 to 3 years in both cohorts, categorical risk classifications did not always correspond to predicted or observed probabilities. For example, the Nagashima score classified patients as low risk despite predicted recurrence probabilities ranging from 25% to 75% at 1 to 3 years, respectively (**Fig. 3**), favouring a probabilistic approach.

**Fig. 3.**
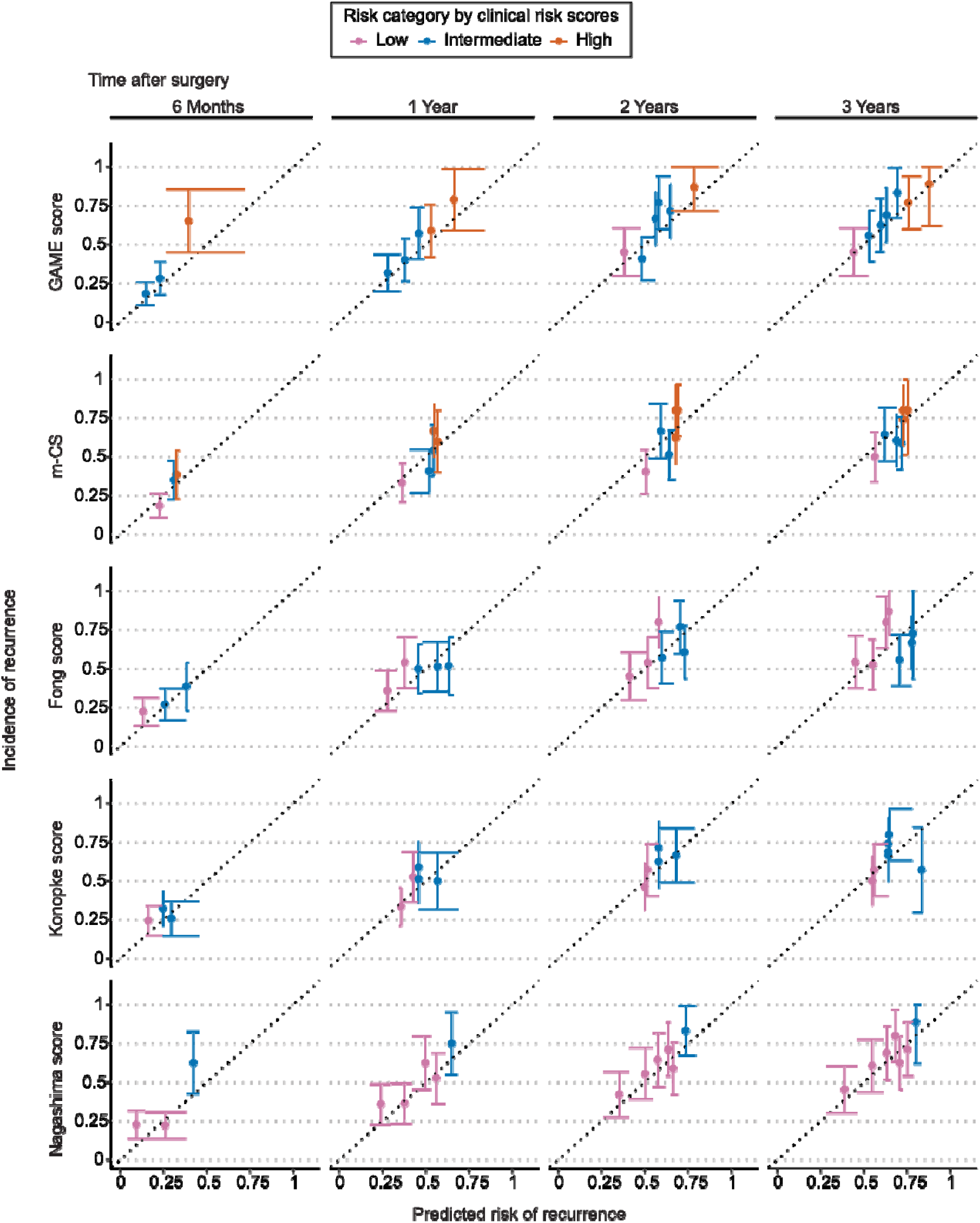
Validation of Recalibrated Clinical Risk Scores in the OSLO-COMET Cohort. Each dot represents the mean predicted risk from models recalibrated on KaroLiver data versus observed recurrence risk in the OSLO-COMET cohort after colorectal cancer liver metastasis resection. Error bars represent 95% confidence intervals, and diagonal dotted lines represent perfect calibration. GAME, genetic and morphological evaluation; m-CS, modified clinical risk score.

Two scores incorporated postoperative variables, including resection margin (Nordlinger) and maximum tumour viability, as well as vascular and biliary invasion (Helsinki). Comparing the performance of these scores with and without postoperative variables (**Table S3**, **Fig. S11**) revealed slightly lower performances (mean ΔAUC at 6 months to 3 years +0.02 [95% CI -0.001 to +0.05] for Helsinki and +0.01 [95% CI - 0.0006 to -0.02] for Nordlinger).

### Clinical Utility of the Recalibrated Models

In KaroLiver, most recalibrated scores showed higher net benefit than default strategies that would assume uniform high or low risk for all patients (**Fig. 4**), with the GAME score yielding the highest net benefit across most time points and threshold probabilities. The m-CS score provided the least incremental benefit. Results in the OSLO-COMET validation cohort were concordant.

**Fig. 4.**
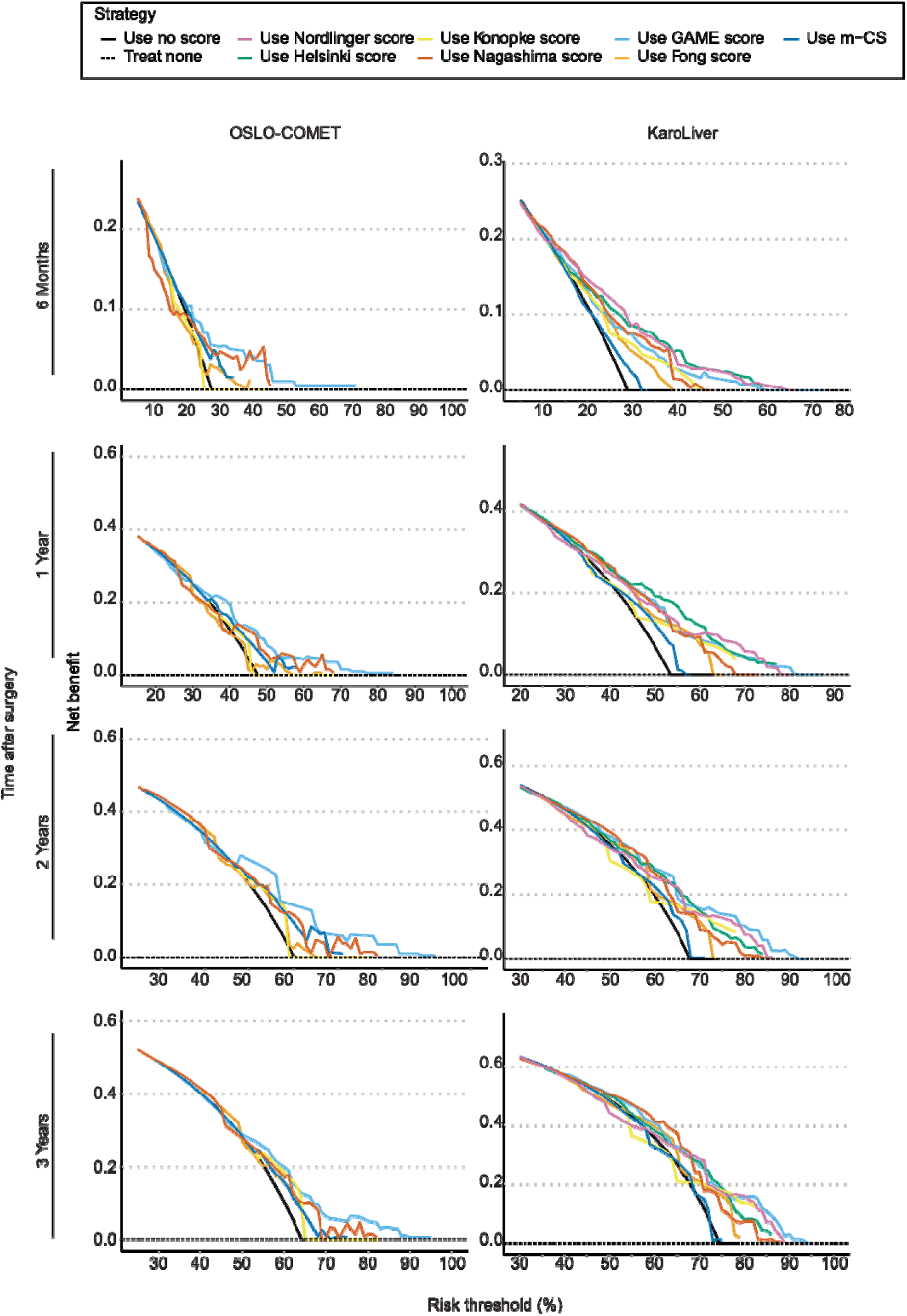
Decision Curve Analysis of Recalibrated Clinical Risk Score Models. A higher net benefit than the “treat all” scenario indicates a better balance between true and false positives with the scoring model. The values have been optimism-corrected using bootstrap internal validation in KaroLiver. The x-axis represents the threshold probability for treatment. CRLM, colorectal cancer liver metastasis; GAME, genetic and morphological evaluation; m-CS, modifie clinical risk score.

### Development of an Online Interface for CRLM Risk Assessment

The regression model coefficients derived from score recalibration (**Table S4**) were used to develop PLATO (**P**ersonalised **L**iver metastases risk **A**ssessmen**T** in **O**ncology; www.plato-calc.org), an open-source calculator for individualised recurrence risk estimation after CRLM resection. PLATO accepts input for the variables required by each validated score, and returns individualised recurrence probabilities at 1, 2, and 3 years after resection, along with the corresponding categorical risk classification from each original score. Its source code, including score calculation, recalibration, and validation pipelines, is publicly available (https://github.com/vilja-a/plato-crlm-risk-prediction/), enabling independent verification, adaptation, and integration of future risk scores.

## Discussion

This study evaluated seven established CRLM risk scores, made the model parameters openly available, and developed PLATO, an online tool that provides individualised recurrence probabilities. External validation supported the generalisability of the probabilistic prediction approach. A central contribution is the conversion of categorical risk strata into percentage-based estimates for all scores, making validated prognostic evidence clinically actionable and enabling direct benchmarking of novel scores. The published coefficients and source code provide an adaptable reference framework for future score calibration, validation, and comparison.

KaroLiver is comparable to other contemporary cohorts with respect to median age, sex distributions, surgical approaches, and molecular profile^38–41^. Its 5-year OS is consistent with the LiverMetSurvey benchmark^42^. Successful validation in OSLO-COMET, a trial cohort with substantially different inclusion criteria, supports the robustness and generalisability of the probability estimates.

All scores significantly stratified RFS, confirming their discriminative value in both cohorts, although performance varied across scores and follow-up intervals. The clinical value of a score depends on the decision it supports, and the meaningful threshold range differs accordingly; intensified surveillance favours lower risk thresholds at earlier time points, where the cost of missing recurrences outweighs that of unnecessary imaging. Adjuvant therapy selection warrants higher thresholds at later time points, where treatment burden must be proportionate to recurrence risk. Hence, probability estimates are most informative when the decision context is defined, and future score/model development would benefit from clinically motivated, *a priori* defined performance targets, tailored to each scenario. Decision curve analysis demonstrated that the GAME score had the highest net benefit at 1 and 2 years. Score performance at lower thresholds and earlier time points remained limited across all scores, highlighting an unmet need that future purpose-built models could address; accordingly, the 6-month time point was not included in PLATO.

To our knowledge, this study provides the first validation of the Helsinki score, which incorporates postoperative histological variables. Adding these histological variables to the Helsinki and Nordlinger scores did not meaningfully improve discrimination, suggesting that classical histological parameters provide limited independent information beyond conventional clinical data in this setting.

The choice of outcome endpoint warrants reflection. There is limited correlation between RFS and OS after CRLM resection, likely due to distinct biological and treatment-related determinants of each outcome^33^. Disease trajectories after CRLM resection are heterogeneous and include liver-only, lung-only, and mixed recurrences that might be amenable to curative-intent treatment. This heterogeneity complicates the use of OS as an endpoint because of increasingly complex and effective post-recurrence treatments. Against this background, the clinical rationale for using RFS as the prognostic target becomes stronger, since RFS more directly reflects tumour biology rather than the variable downstream treatment course. Beyond binary recurrence, the concept of “treatable recurrence,” amenable to curative-intent retreatment, represents a clinically relevant refinement not yet captured by current scores. Models targeting early extrahepatic recurrence partly address this^43^, but scoring systems have yet to integrate recurrence patterns into individualised probability estimates^43^.

A recent study compared multiple risk scores and, in line with our results, demonstrated that most established scores significantly stratified patients by OS, although discrimination was modest across models^44^. The study provided a valuable head-to-head comparison of established scores; however, calibration, net benefit, external validation, and translation into individualised probability estimates were beyond its scope. Previous calculators have advanced individualised prognostication^45–50^, but several limitations restrict direct benchmarking and clinical translation: reliance on machine learning models with limited parameter transparency^45,46^, absent external validation, unavailability of model parameters, restriction to selected patient subgroups^48^, and focus on long-term outcomes^50^. We address these limitations by providing individualised probability estimates at clinically relevant time points, grounded in established scores with documented validation histories, and by publishing source code and model coefficients.

Next-generation clinical risk scores will increasingly incorporate molecular tumour profiling, including sequencing, imaging-based machine learning, and other high-dimensional data. As clinical decision support moves toward AI-based systems^51^, validated and interpretable risk scores remain essential: broad clinical trust in AI-generated recommendations requires transparent, externally validated benchmark models against which their outputs can be contextualised. By publishing open model coefficients alongside calibration and decision-curve analyses, we aim to provide a transparent, reproducible reference against which such future prognostic tools can be evaluated, as treatment options and prognostic variables evolve.

While this study builds on the established clinical risk scores, offering extensive evaluation, a transparent workflow and improved accessibility via PLATO, several limitations are worth noting. First, both cohorts consist predominantly of patients with European ancestry treated within publicly funded health systems, which limits generalisability to populations with differing disease biology, treatment access, or healthcare structures. Second, genetic variant data were available for 44% of KaroLiver patients, necessitating imputation and potentially affecting the robustness of the m-CS and GAME scores. Finally, differences in postoperative surveillance intervals between cohorts, every 3 to 6 months for KaroLiver and every 4 months for OSLO-COMET, may have introduced bias in the timing of recurrence detection. Despite these limitations, our findings provide a transparent foundation for future prognostic score development and benchmarking with openly available model parameters. Properly recalibrated and externally validated, these established clinical risk scores provide the reference frame against which next-generation prognostic models can be measured – and PLATO makes that comparison clinically accessible.

## Data Availability

Pseudonymised clinical data from the KaroLiver and OSLO-COMET cohorts are not publicly available because of legal and ethical restrictions on patient data. The corresponding authors (M.G., J.E.) will share KaroLiver data upon reasonable request, contingent on approval by the Swedish Ethical Review Authority and execution of a research collaboration/data transfer agreement.

https://github.com/vilja-a/plato-crlm-risk-prediction/

https://plato-calc.org/app/plato-crlm-risk-prediction

## Previous Communication

This work has not been presented previously at a society meeting or conference.

## Acknowledgements

Karolinska Institutet (KI) supports KaroLiver as a resource through the KI Microenvironmental Control of Metastases network, which received funding from the Board of Research at KI. The study was supported by The Swedish Research Council (projects nr. 2018-02023 and 2024-03026, to M.G.), StratRegen (to M.G.), The Swedish Society for Medical Research (to M.G.), The Swedish Cancer Society (22 2175 Pj and 25 4754 Pj to M.G.), the Center for Innovative Medicine (to M.G. and J.E.), and Radiumhemmet’s research funds (to M.G. and J.E). JE was supported by Region Stockholm and the Bengt Ihre Foundation. N.G. was supported by the German Research Foundation (#460567311) and Ruth and Richard Julin’s foundation. This study was not preregistered.

The authors acknowledge the use of OpenAI’s ChatGPT (various model versions available through the ChatGPT platform, OpenAI, June 2024–July 2026) and Anthropic’s Claude (Claude Sonnet 4.6, Anthropic, 2026) during the preparation of this work. These tools were used to assist with code writing, debugging, error detection, and text formulation. All AI-generated outputs were critically reviewed and verified by the authors, who take full responsibility for the final content.

The authors would like to thank SciLifeLab Serve for hosting the PLATO online tool. The authors declare no financial disclosures or conflicts of interest relevant to this work.

## Code Availability

Data processing and analysis code, including the PLATO calculator, is publicly available on GitHub at the URL provided in the Methods section.

