## supplementary_material for "Individualised and Time-Dependent Prediction of Recurrence After Colorectal Cancer Liver Metastases Resection"

*equal contribution, corresponding authors

**Corresponding authors**

C1:77

14186 Huddinge, Sweden

**Supplementary Materials - Index**

| **Supplementary Methods** |  |
| --- | --- |
| Full Description of Statistical Analyses and Online Tool Development | *pag. 3* |
| **Supplementary Figures and Tables** |  |
| **Table S1.** TRIPOD+AI checklist | *pag. 7* |
| **Table S3.** Time-Dependent Area-Under-Curve of Complete Scoring Models and Limited Models without Postoperative Factors | *pag. 12* |
| **Fig. S1.** Flowchart of Patient Selection | *pag. 13* |
| **Fig. S2.** Quality Control of Imputation in KaroLiver | *pag. 14* |
| **Fig. S3.** Quality Control of Imputation in OSLO-COMET | *pag. 15* |
| **Fig. S4.** Overall Survival Stratified by Clinical Risk Scores in KaroLiver | *pag. 16* |
| **Fig. S5.** Multivariable Analyses of Fong, m-CS, GAME, and Nagashima Score Risk Factors in KaroLiver (Recurrence-Free Survival) | *pag. 17* |
| **Fig. S6.** Multivariable Analyses of Konopke, Helsinki, and Nordlinger Score Risk Factors in KaroLiver (Recurrence-Free Survival) | *pag. 18* |
| **Fig. S7.** Multivariable Analyses of Fong, m-CS, GAME, and Nagashima Score Risk Factors in KaroLiver (Overall Survival) | *pag. 19* |
| **Fig. S8.** Multivariable Analyses of Konopke, Helsinki, and Nordlinger Score Risk Factors in KaroLiver (Overall Survival) | *pag. 20* |
| **Fig. S9.** Calibration of Recalibrated Clinical Risk Score Models in KaroLiver | *pag. 21* |
| **Fig. S10.** Calibration of Recalibrated Clinical Risk Score Models After Bootstrap Internal Validation in KaroLiver | *pag. 22* |
| **Fig. S11.** Decision Curve Analysis for Recalibrated Postoperative Scoring Models with and without Postoperative Factors in KaroLiver | *pag. 23* |
| **References** | *pag. 24* |

**Supplementary Methods** Full Description of Statistical Analyses and Online Tool Development

**Data Management and Visualisation**

All statistical analyses were performed in R v 4.4.^1^ via Rstudio IDE v 2024.4.0^2^. We used R packages tidyr v 1.3^3^, tidyverse v 2.0^4^, dplyr v 1.1^5^ for general data management and preprocessing. Data visualisation was done with ggplot2 3.5^6^, ggthemes 5.1^7^ and ggpubr v 0.6^8^. Summary tables of clinical cohort were created with gtsummary v 2.0^9^.

**Imputation of Missing Data**

We used Multiple Imputation by Chained Equations (MICE) to handle missing data with default settings, except by manually adjusting the imputation formula to include the baseline hazard for recurrence-free survival, resulting in m = 5 imputed datasets. In KaroLiver, clinical variables including the presence of simultaneous lung metastases, radiological measure of the largest liver metastasis size (mm), primary tumour T and N stages, serum CEA levels (µg/L), *RAS*-mutational status, the presence of vascular or biliary invasions, resection margin (mm), and tumour viability (%) had missing observations that were imputed (**Fig. S2A**). In OSLO-COMET, missing observations were imputed for clinical variables of radiological measures of the largest liver metastasis size (mm), resection margin (mm), primary tumour T and N stages, serum CEA levels (µg/L), *RAS*-mutational status, and the presence of extrahepatic metastases (**Fig. S3A**). We used predictive mean matching for continuous variables, logistic regression for binary variables, and polytomous logistic regression for unordered categorical data.

**Quality Control and Evaluation After Data Imputation**

For each imputed variable, its distribution within the imputed data sets was compared to the original data set. Continuous variables were evaluated by using histograms and descriptive statistics, while binary or ordinal variables were assessed using category proportions (**Fig. S2B-D & S3B-D**). From these alternative data sets, we selected the one showing the highest statistical similarity: set #2 for KaroLiver and #3 for OSLO-COMET.

**Statistical Testing**

All statistical testing was performed with rstatix v 0.7^11^ or built-in functions of base R. Nonparametric Wilcoxon rank-sum tests were used to compare the distributions of continuous clinical variables, and the paired form to compare the distributions between the original and imputed data sets. For binary clinical variables, the distributions were evaluated using pairwise McNemar tests, whereas for ordinal variables, paired chi-squared tests were applied.

When evaluating the difference between the clinical characteristics of KaroLiver and OSLO-COMET, a chi-squared test was applied in the case of binary variables, and a Wilcoxon rank sum test in the case of ordinal variables. Benjamini-Hochberg multiple testing correction was used to control the false discovery rate.

**Computation of Clinical Risk Scores**

To externally validate Helsinki^12^, modified clinical risk score (m-CS)^13^, Fong^14^, Nordlinger^15^, Nagashima^16^, Konopke^17^, and Genetic and Morphological Evaluation (GAME)^18^ scores, we followed the TRIPOD statement for external validation for clinical risk scores (**Table S1**)^19^. Patients were grouped into categories of low (Fong 0–2, GAME 0–1, Konopke 1, m-CS 0, Nagashima Grade A), intermediate (Fong 3–4, GAME 2–3, Konopke 2, m-CS 1, Nagashima Grade B), high (Fong 5, GAME 4–7, Konopke 3, m-CS 2, Nagashima grade C), and very high (m-CS 4) risk for CRLM recurrence according to each clinical score description.

As part of the GAME score, tumour burden score (TBS)^20^ was determined using the following formula:

$$TBS= \sqrt{d_{\max}^{2}+n_{\mathrm{met}}^{2}}$$

where

d_max_ = radiological measure for maximum diameter (cm) of liver metastasis

n_met_ = radiological measure for the number of metastases in the liver

**Evaluation of Competing Risks**

We assessed death as a competing risk by comparing cumulative incidence with and without adjustment for competing risks using cmprsk v 2.2. Death contributed 0.2-6% to cumulative incidence over 10 years, indicating a minor influence on the relapse as an event. Furthermore, only 6.3% (n=45) of deaths before CRLM recurrence during the first 3 years were present in the dataset; therefore, we deemed it unnecessary to adjust for death as a competing risk.

**Recalibration of the Clinical Risk Scores**

To evaluate each score’s performance in CRLM recurrence risk prediction, logistic regression models were fitted to the outcome variable (i.e., recurrence) at 6 months and 1, 2, and 3 years after CRLM resection, using risk factors computed for each clinical risk score in the KaroLiver cohort. The continuous Nagashima score was modelled using restricted cubic splines to provide greater flexibility, and categorical variables present in the other clinical risk scores were entered as indicator variables. To fulfil the criteria of a fully observed outcome variable, patients who were lost to follow-up (n=28, 3.9%) at 3 years were excluded before fitting the models for 3-year risk prediction (**Fig. S1**). Outcome was fully observed for the other time points of risk evaluation. Package rms v 6.8^22^ was used for model fitting:

$$\mathrm{logit}\left( P\left( Y=1 \right) \right)=log\left( \frac{P(Y=1)}{1-P\left( Y=1 \right)} \right)=X\hat{\beta}$$

The resulting models were used to predict relapse risk P for each observation using the predicted probability formula:

$$P\left( Y=1 \right)= \frac{1}{1+e^{-(X\hat{\beta})}}$$

Linear predictor:

$$X\hat{\beta}=\beta_{o}$$

$$+\sum_{j=1}^{q} \gamma_{j}\cdot\left[ c_{j} \right]$$

$$+ \beta_{1}x+\sum_{k=1}^{K-2} \delta_{k}{(x-t_{k})}_{+}^{3}$$

Where

$$\left[ c_{j} \right]=\left\{ \begin{aligned} 1 \text{if subject is in group}c_{j} \\ 0 \text{otherwise } \end{aligned} \right.$$

$${(x-t_{k})}_{+}^{3}= \left\{ \begin{aligned} {(x-t_{k})}^{3} if x>t_{k} \\ 0 otherwise \end{aligned} \right.$$

P is the probability for CRLM relapse at 6 months, 1 year, 2 years or 3 years after operation

b_0_: intercept

g: coefficient for group c_j_, j= 1, …,q

b_1_: linear coefficient for continuous predictor x

d_k_: spline coefficient at knot t_k_, k=1, …, K-2

t_1_ < … <t_K_: knot locations (K knots total)

The model coefficients for all clinical risk scores across the evaluated timepoints are shown in **Table S4**.

**Model Performance Analyses**

We evaluated the clinical risk scores’ performance at predicting CRLM relapse after resection by examining model discrimination, calibration and net benefit following the framework for development and validation of clinical prediction models^23^. For internal validation of the recalibrated models, bootstrap resampling with 500 bootstrap samples was added to estimate the effect of overfitting with calibration and net benefit analyses in KaroLiver. For external validation of the recalibrated models, model coefficients were used to compute the risk estimates in OSLO-COMET with probability formula. To examine the added value of postoperative parameters part of Helsinki and Nordlinger scores, the full model of these scores were compared to the limited version of the models including only preoperative variables in KaroLiver.

**Discrimination**

The time-dependent area under the ROC curve approach was used to evaluate the discrimination ability of the prognostic scores at multiple time points. Unlike the conventional ROC curve approach, it accounts for censoring and changes in event status over time.^24^ Therefore time-dependent ROC curves were used to evaluate discrimination over time with timeROC v 0.4^24^. Survival analysis in evaluating the clinical risk scores’ capability to discriminate between the overall and recurrence-free survival across risk classes of each clinical risk score. For the purpose, we used Kaplan-Meier and multivariable analysis of all scoring factors with R packages survminer v 0.5^25^, survival v 3.7^26^ Evaluation of discrimination did not require model recalibration, and therefore, it was only implemented in KaroLiver.

**Calibration**

Calibration of each risk score was evaluated by comparing the expected outcomes from the recalibrated models with the observed outcomes. Individuals in the study cohort were ordered in increasing order by predicted risk from each clinical risk score, and the data were divided into 8-12 groups with equal numbers of recurrences in each group; in KaroLiver, 30-40; in OSLO-COMET, 20 across all timepoints. Cumulative incidence within these subgroups, with 95% confidence intervals, was evaluated using cmprsk v 2.2^21^. The result was visualised using calibration plots, with the predicted mean risk plotted against the mean cumulative incidence.

**Net Benefit**

Net benefit NB(p_t_) across threshold probabilities p_t_ between 0 and 1 was computed with the following formula by Vickers et al.^27^:

$$NB\left( p_{t} \right)=\frac{True Positives}{N}- \frac{False Positives}{N}\left( \frac{p_{t}}{1-p_{t}} \right)$$

$$= \frac{I\left( T=t_{x}|p\geq p_{t} \right)\times m}{N}-\frac{\left( 1-I\left( T=t_{x}|p\geq p_{t} \right)\times m \right)}{N}\left( \frac{p_{t}}{1-p_{t}} \right)$$

Where

I = cumulative incidence

t_x_ = time point of interest after operation

p= predicted risk by the clinical risk score

p_t_= threshold probability

m=number of true positives (individuals with predicted risk p³p_t_)

N= total number of individuals

Cumulative incidence calculated with cmprsk v 2.2^21^ was used to assess true positive values of relapses at the time point of interest and compared to the predicted relapse risk based on the clinical risk scores at the given time point of interest.

**Development of an Online Prediction Tool for CRLM Recurrence**

Model coefficients from the recalibration of the scores (**Table S4**) were used to build the online application PLATO with Shiny v 1.11.^28^ The application allows the user to calculate the recurrence risk for an individual patient using the probability formula, selecting the risk parameters from the online interface. PLATO uses the model coefficients interactively with user input to calculate the recurrence risk at 1, 2, and 3 years after CRLM resection based on each of the 7 clinical risk scores, and in addition to the percentage of relapse risk, it gives the original score value and the risk group based on the score. The user can also explore the actual recurrences within KaroLiver grouped by the original clinical risk score groups.

**Code Availability**

The code for model fitting by bootstrapping, discrimination, calibration and net benefit used in this study was done as previously described^23,29^, with modifications to accommodate the current study design. All the code to reproduce the analyses and build the Shiny interface has been published at the GitHub repository https://github.com/vilja-a/plato-crlm-risk-prediction/.

**Supplementary Figures and Tables**

**Table S1.** TRIPOD+AI checklist From: Collins GS, Moons KGM, Dhiman P, et al. *BMJ* 2024;385:e078378. doi:10.1136/bmj-2023-078378

| **Section/**  **Topic** | **Item** | **Development/ evaluation*** | **Checklist item** | **Reported on page** |
| --- | --- | --- | --- | --- |
| **TITLE** | | | | |
| *Title* | 1 | D;E | Identify the study as developing or evaluating the performance of a multivariable prediction model, the target population, and the outcome to be predicted | 1 |
| **ABSTRACT** | | | | |
| *Abstract* | 2 | D;E | See TRIPOD+AI for Abstracts checklist | 3 |
| **INTRODUCTION** | | | | |
| *Background* | 3a | D;E | Explain the healthcare context (including whether diagnostic or prognostic) and rationale for developing or evaluating the prediction model, including references to existing models | 4-5 |
|  | 3b | D;E | Describe the target population and the intended purpose of the prediction model in the context of the care pathway, including its intended users (e.g., healthcare professionals, patients, public) | 4-5 |
|  | 3c | D;E | Describe any known health inequalities between sociodemographic groups | NA |
| *Objectives* | 4 | D;E | Specify the study objectives, including whether the study describes the development or validation of a prediction model (or both) | 5 |
| **METHODS** | | | | |
| *Data* | 5a | D;E | Describe the sources of data separately for the development and evaluation datasets (e.g., randomised trial, cohort, routine care or registry data), the rationale for using these data, and representativeness of the data | 6-7 |
|  | 5b | D;E | Specify the dates of the collected participant data, including start and end of participant accrual; and, if applicable, end of follow-up | 6-7 |
| *Participants* | 6a | D;E | Specify key elements of the study setting (e.g., primary care, secondary care, general population)  including the number and location of centres | 6 |
|  | 6b | D;E | Describe the eligibility criteria for study participants | 6-7 |
|  | 6c | D;E | Give details of any treatments received, and how they were handled during model development or evaluation, if relevant | 7-8 |
| *Data preparation* | 7 | D;E | Describe any data pre-processing and quality checking, including whether this was similar across  relevant sociodemographic groups | 8 |
| *Outcome* | 8a | D;E | Clearly define the outcome that is being predicted and the time horizon, including how and when assessed, the rationale for choosing this outcome, and whether the method of outcome assessment is consistent across sociodemographic groups | 8 |
|  | 8b | D;E | If outcome assessment requires subjective interpretation, describe the qualifications and demographic characteristics of the outcome assessors | 8 |
|  | 8c | D;E | Report any actions to blind assessment of the outcome to be predicted | NA |
| *Predictors* | 9a | D | Describe the choice of initial predictors (e.g., literature, previous models, all available predictors) and any pre-selection of predictors before model building | 8 |
|  | 9b | D;E | Clearly define all predictors, including how and when they were measured (and any actions to blind assessment of predictors for the outcome and other predictors) | 8-9 |
|  | 9c | D;E | If predictor measurement requires subjective interpretation, describe the qualifications and demographic characteristics of the predictor assessors | NA |
| *Sample size* | 10 | D;E | Explain how the study size was arrived at (separately for development and evaluation), and justify that the study size was sufficient to answer the research question. Include details of any sample size calculation | 6-7,  Fig. S1,  Supplementary Methods |
| *Missing data* | 11 | D;E | Describe how missing data were handled. Provide reasons for omitting any data | 8-9, Supplementary Methods |
| *Analytical methods* | 12a | D | Describe how the data were used (e.g., for development and evaluation of model performance) in the analysis, including whether the data were partitioned, considering any sample size requirements | 8-9, Supplementary Methods |
|  | 12b | D | Depending on the type of model, describe how predictors were handled in the analyses (functional form,  rescaling, transformation, or any standardisation). | 9, Supplementary Methods |
|  | 12c | D | Specify the type of model, rationale^†^, all model-building steps, including any hyperparameter tuning, and method for internal validation | 9, Supplementary Methods |
|  | 12d | D;E | Describe if and how any heterogeneity in estimates of model parameter values and model performance was handled and quantified across clusters (e.g., hospitals, countries). See TRIPOD-Cluster for additional considerations^‡^ | NA |
|  | 12e | D;E | Specify all measures and plots used (and their rationale) to evaluate model performance (e.g., discrimination, calibration, clinical utility) and, if relevant, to compare multiple models | 8-9, Supplementary Methods |
|  | 12f | E | Describe any model updating (e.g., recalibration) arising from the model evaluation, either overall or for particular sociodemographic groups or settings | 9, Supplementary Methods |
|  | 12g | E | For model evaluation, describe how the model predictions were calculated (e.g., formula, code, object, application programming interface) | 9, Supplementary Methods |
| *Class imbalance* | 13 | D;E | If class imbalance methods were used, state why and how this was done, and any subsequent methods to  recalibrate the model or the model predictions | NA |
| *Fairness* | 14 | D;E | Describe any approaches that were used to address model fairness and their rationale | NA |
| *Model output* | 15 | D | Specify the output of the prediction model (e.g., probabilities, classification). Provide details and rationale for any classification and how the thresholds were identified | 9 |
| *Training versus*  *evaluation* | 16 | D;E | Identify any differences between the development and evaluation data in healthcare setting, eligibility criteria, outcome, and predictors | 6-7, 10, Table 1 |
| *Ethical approval* | 17 | D;E | Name the institutional research board or ethics committee that approved the study and describe the participant-informed consent or the ethics committee waiver of informed consent | 6 |
| **OPEN SCIENCE** | | | | |
| *Funding* | 18a | D;E | Give the source of funding and the role of the funders for the present study | 2,18 |
| *Conflicts of interest* | 18b | D;E | Declare any conflicts of interest and financial disclosures for all authors | 2,19 |
| *Protocol* | 18c | D;E | Indicate where the study protocol can be accessed or state that a protocol was not prepared | Not prepared |
| *Registration* | 18d | D;E | Provide registration information for the study, including register name and registration number, or state  that the study was not registered | Not registered |
| *Data sharing* | 18e | D;E | Provide details of the availability of the study data | 2 |
| *Code sharing* | 18f | D;E | Provide details of the availability of the analytical code^§^ | 9, 14, 19, Supplementary Methods |
| **PATIENT & PUBLIC INVOLVEMENT** | | | | |
| *Patient & Public Involvement* | 19 | D;E | Provide details of any patient and public involvement during the design, conduct, reporting, interpretation, or dissemination of the study or state no involvement. | No involvement |
| **RESULTS** | | | | |
| *Participants* | 20a | D;E | Describe the flow of participants through the study, including the number of participants with and without the outcome and, if applicable, a summary of the follow-up time. A diagram may be helpful. | 6-7, 10, Fig. S1 |
|  | 20b | D;E | Report the characteristics overall and, where applicable, for each data source or setting, including the key dates, key predictors (including demographics), treatments received, sample size, number of outcome events, follow-up time, and amount of missing data. A table may be helpful. Report any  differences across key demographic groups. | 10-11, Table 1, Fig. 1b |
|  | 20c | E | For model evaluation, show a comparison with the development data of the distribution of important predictors (demographics, predictors, and outcome). | 10, Table 1, Fig. 1b |
| *Model development* | 21 | D;E | Specify the number of participants and outcome events in each analysis (e.g., for model development, hyperparameter tuning, model evaluation) | 10, 13, Supplementary Methods, Fig. S1 |
| *Model specification* | 22 | D | Provide details of the full prediction model (e.g., formula, code, object, application programming interface) to allow predictions in new individuals and to enable third-party evaluation and implementation, including any restrictions to access or re-use (e.g., freely available, proprietary)^‖^ | 9, Supplementary Methods, Table S4 |
| *Model performance* | 23a | D;E | Report model performance estimates with confidence intervals, including for any key subgroups (e.g., sociodemographic). Consider plots to aid presentation. | 12-14, Fig. 3&4, S9&S10, Table S2 |
|  | 23b | D;E | If examined, report results of any heterogeneity in model performance across clusters. See TRIPOD  Cluster for additional details^‡^. | NA |
| *Model updating* | 24 | E | Report the results from any model updating, including the updated model and subsequent performance | 12-14,  Fig. 3&4, Table S2 |
| **DISCUSSION** | | | | |
| *Interpretation* | 25 | D;E | Give an overall interpretation of the main results, including issues of fairness in the context of the  objectives and previous studies | 15-17 |
| *Limitations* | 26 | D;E | Discuss any limitations of the study (such as a non-representative sample, sample size, overfitting, missing data) and their effects on any biases, statistical uncertainty, and generalizability | 18 |
| *Usability of the model in the context of current care* | 27a | D | Describe how poor quality or unavailable input data (e.g., predictor values) should be assessed and handled when implementing the prediction model | NA |
|  | 27b | D | Specify whether users will be required to interact in the handling of the input data or use of the model, and what level of expertise is required of users | NA |
|  | 27c | D;E | Discuss any next steps for future research, with a specific view to applicability and generalizability of  the model | 17-18 |

*D=items relevant only to the development of a prediction model; E=items relating solely to the evaluation of a prediction model; D;E=items applicable to both the development and evaluation of a prediction model

†Separately for all model building approaches.

‡TRIPOD-Cluster is a checklist of reporting recommendations for studies developing or validating models that explicitly account for clustering or explore heterogeneity in model performance (eg, at different hospitals or centres). Debray et al, BMJ 2023; 380: e071018 [DOI: 10.1136/bmj-2022-071018]

§This relates to the analysis code, for example, any data cleaning, feature engineering, model building, evaluation.

‖This relates to the code to implement the model to get estimates of risk for a new individual.

**Table S3.** Time-Dependent Area-Under-Curve of Complete Scoring Models and Limited Models without Postoperative Factors

|  | **Helsinki Score** | | | **Nordlinger Score** | | |
| --- | --- | --- | --- | --- | --- | --- |
| **Time After CRLM resection Years)** | **AUC (Full Model)** | **AUC (Preoperative Factors)** | **∆AUC** | **AUC (Full Model)** | **AUC  (Preoperative Factors)** | **∆AUC** |
| 0.5 | 0.65 | 0.636 | 0.014 | 0.715 | 0.704 | 0.011 |
| 1 | 0.687 | 0.659 | 0.028 | 0.68 | 0.671 | 0.009 |
| 1.5 | 0.677 | 0.655 | 0.022 | 0.67 | 0.66 | 0.01 |
| 2 | 0.678 | 0.654 | 0.024 | 0.683 | 0.672 | 0.011 |
| 2.5 | 0.679 | 0.641 | 0.038 | 0.679 | 0.666 | 0.013 |
| 3 | 0.693 | 0.66 | 0.033 | 0.702 | 0.688 | 0.014 |

**Fig. S1 Flowchart of Patient Selection**

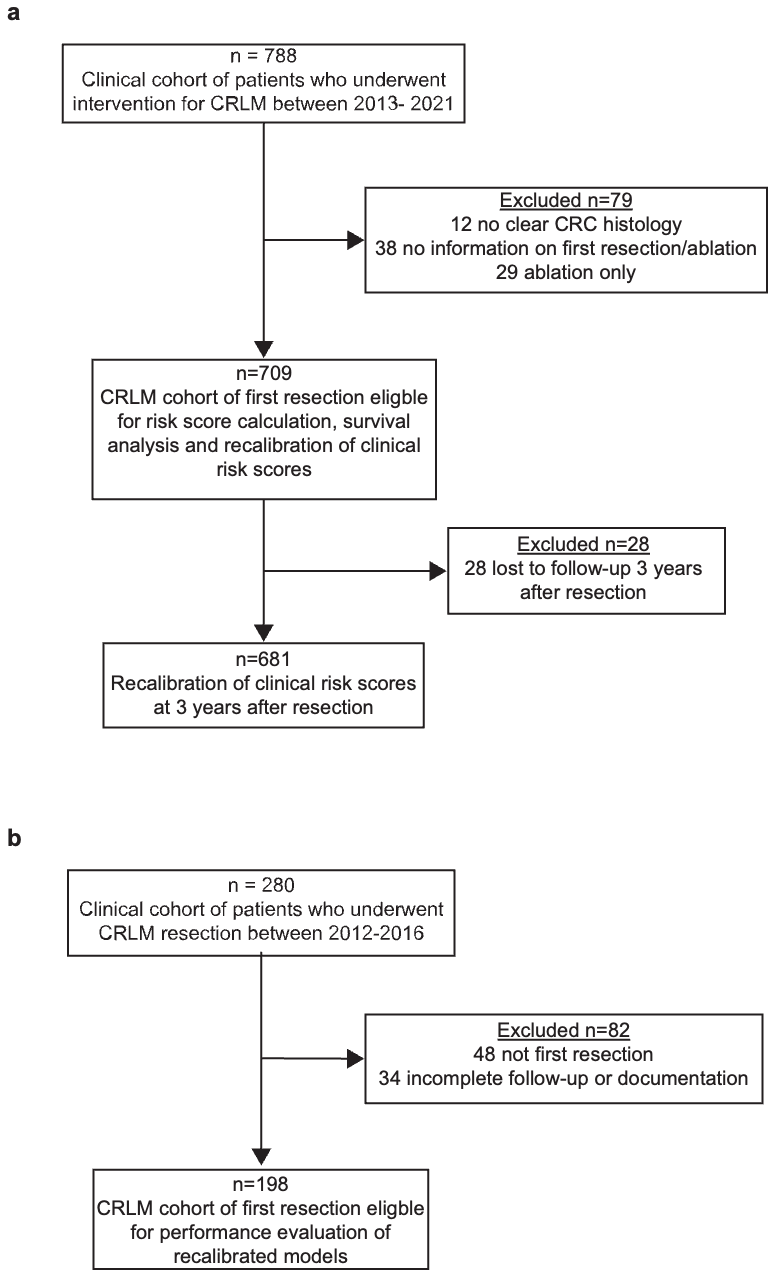

**a** For KaroLiver and **b** OSLO-COMET cohorts. CRLM, colorectal liver metastasis.

**Fig. S2. Quality Control of Imputation in KaroLiver**

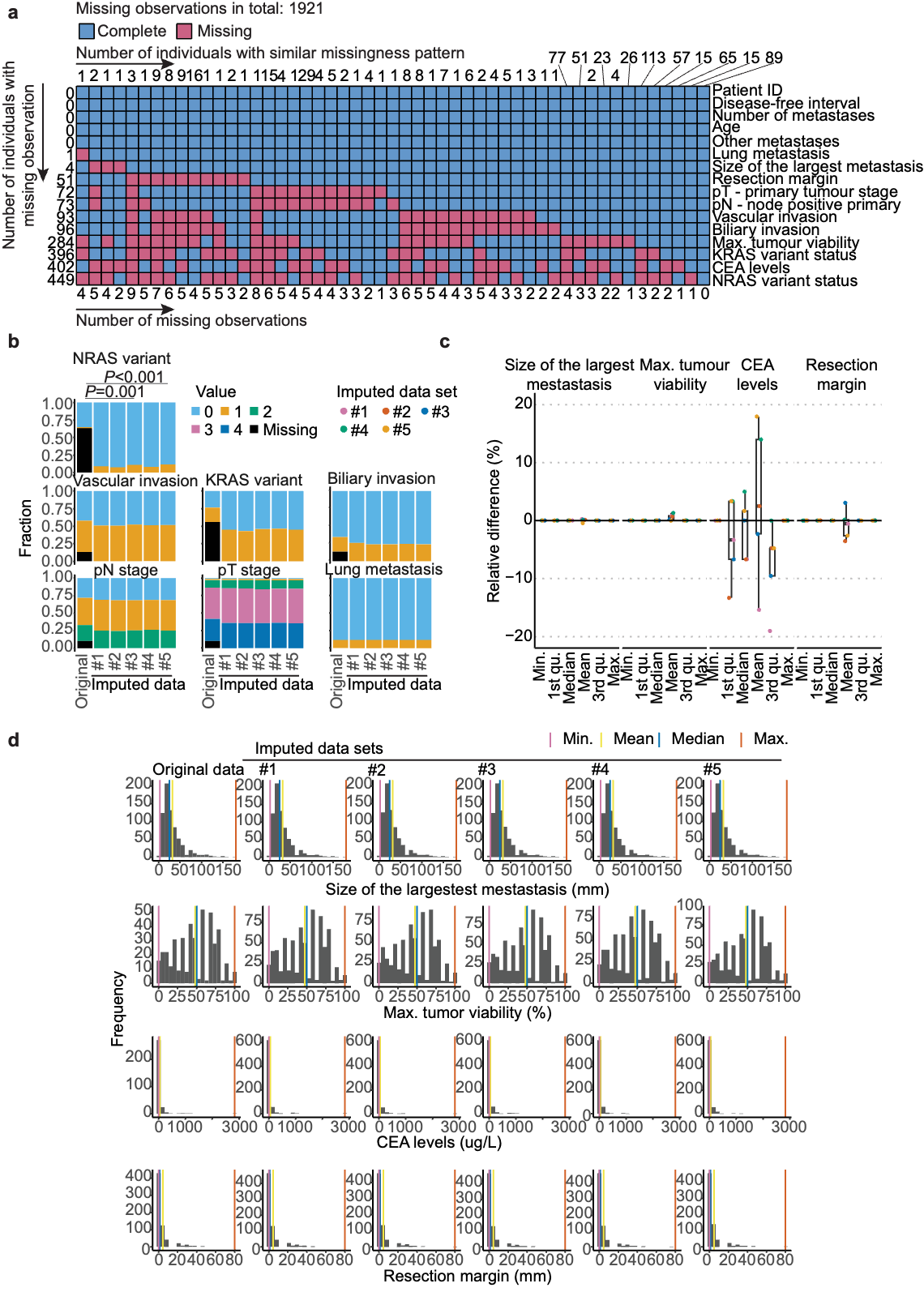

**a** Missingness pattern **b** Distribution of ordinal variables in the original and imputed data sets. Only statistically significant *P* values from McNemar tests are shown; all other comparisons were non-significant. **c** Box-and-whiskers plots of relative differences in descriptive statistics for continuous clinicopathological variables between the original and imputed data sets. Line, median; box, interquartile range (IQR), minimum and maximum values within 1.5xIQR; dots, individual datapoints. **d** Distribution of continuous variables in original and imputed data sets. Statistical comparisons were performed across all panels using appropriate tests. CEA, carcinoembryonic antigen; ID, identifier; Max, maximum; Min, minimum; qu, quartile.

**Figure S3. Quality Control of Imputation in OSLO-COMET**

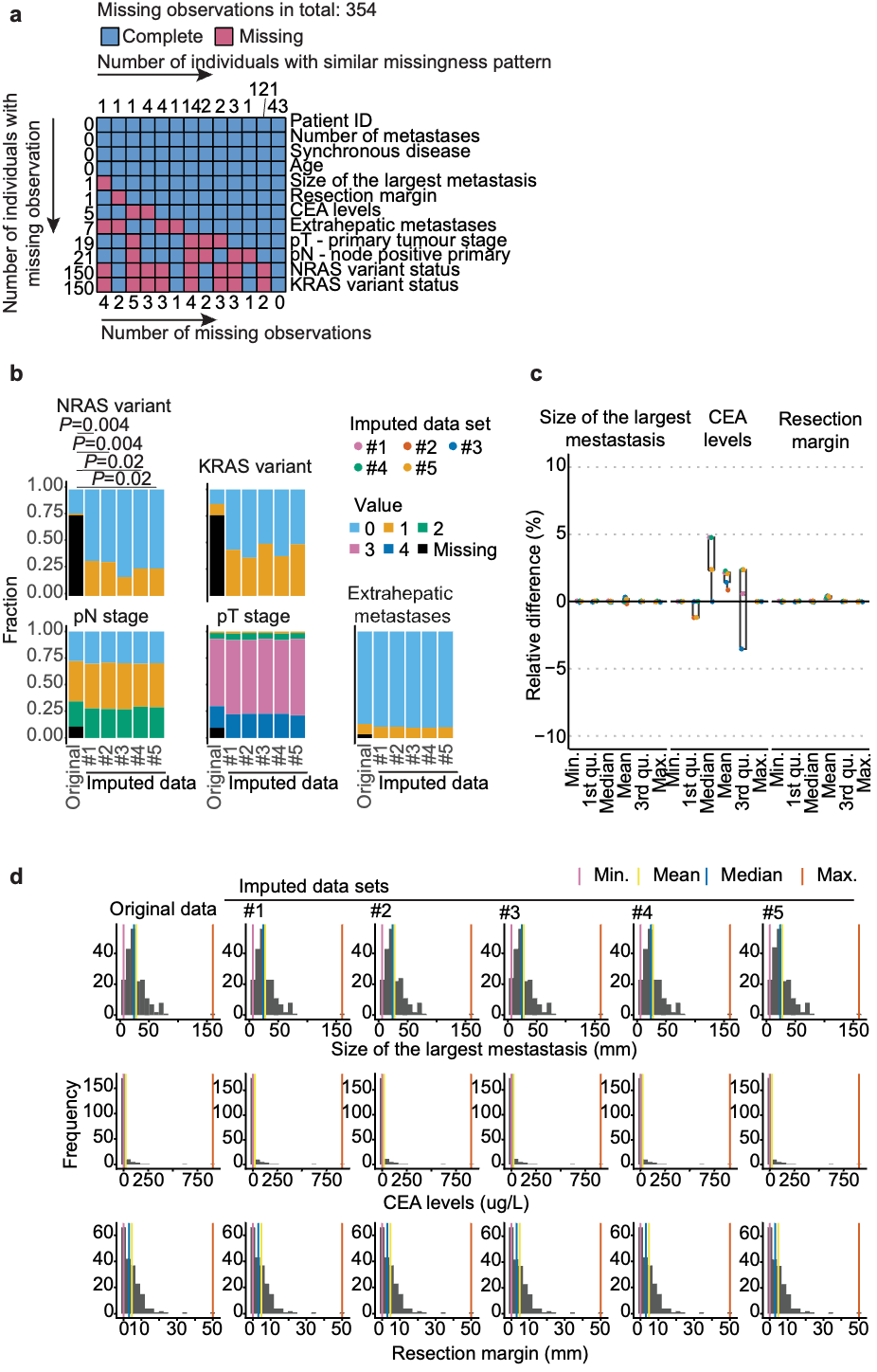

**a** Missingness pattern **b** Distribution of ordinal variables in the original and imputed data sets. Only statistically significant *P* values from McNemar test are shown; all other comparisons were non-significant. **c** Box-and-whiskers plots of relative difference in descriptive statistics for continuous clinicopathological variables between the original and imputed data sets. Line, median; box, interquartile range (IQR), minimum and maximum values within 1.5xIQR; dots, individual datapoints. **d** Distribution of continuous variables in the original and imputed data sets. Statistical comparisons were performed across all panels using appropriate tests. CEA, carcinoembryonic antigen; ID, identifier; Max, maximum; Min, minimum; qu, quartile.

**Fig. S4. Overall Survival Stratified by Clinical Risk Scores in KaroLiver**

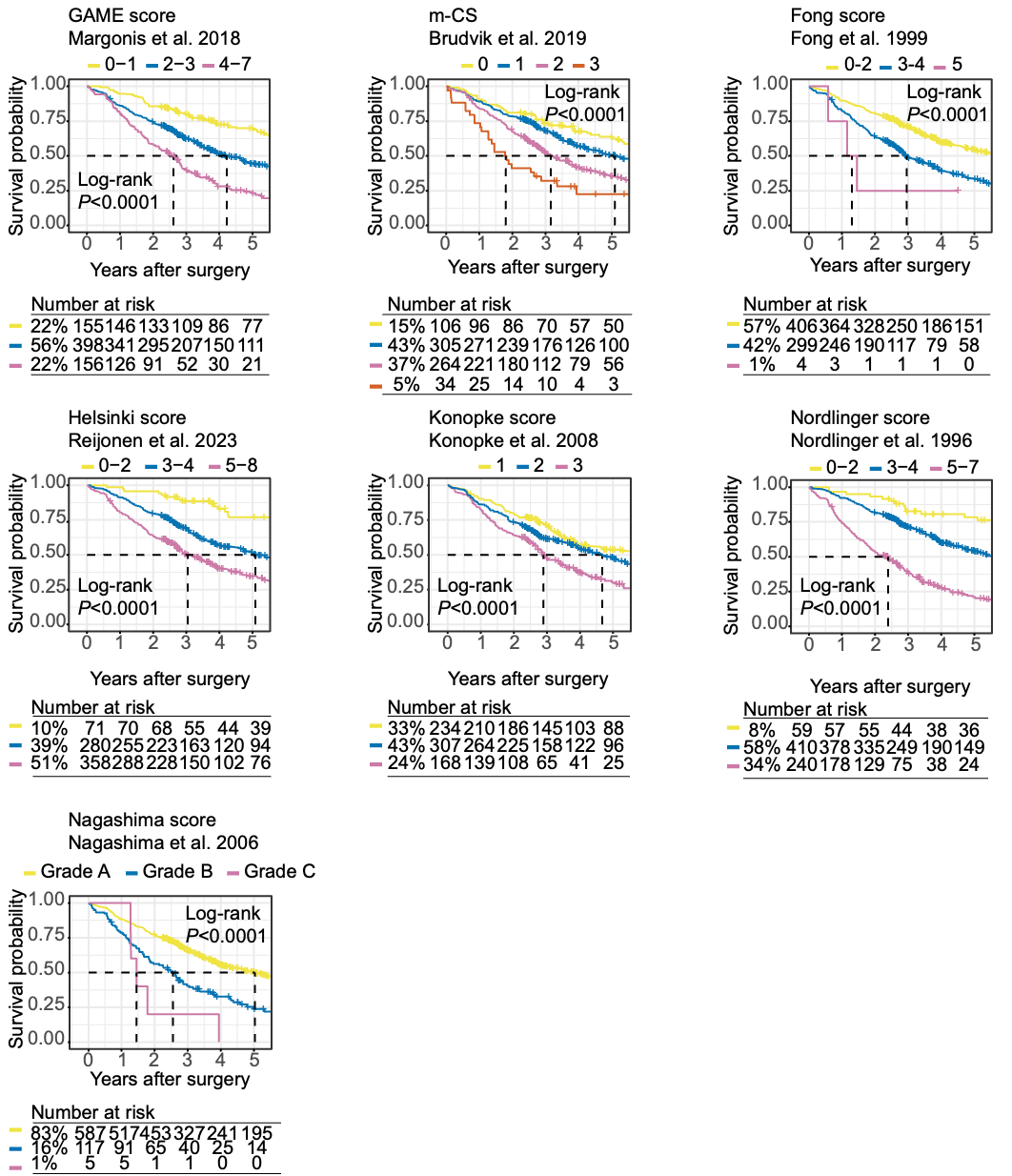

Kaplan-Meier estimates of overall survival stratified as per the original strata of the clinical risk scores. Number at risk, *P* values, statistical tests, and strata definitions are displayed in the plot. CRLM, colorectal cancer liver metastasis; GAME, genetic and morphological evaluation; m-CS: modified clinical risk score.

**Fig. S5 Multivariable Analyses of Fong, m-CS, GAME, and Nagashima Score Risk Factors in KaroLiver (Recurrence-Free Survival)**

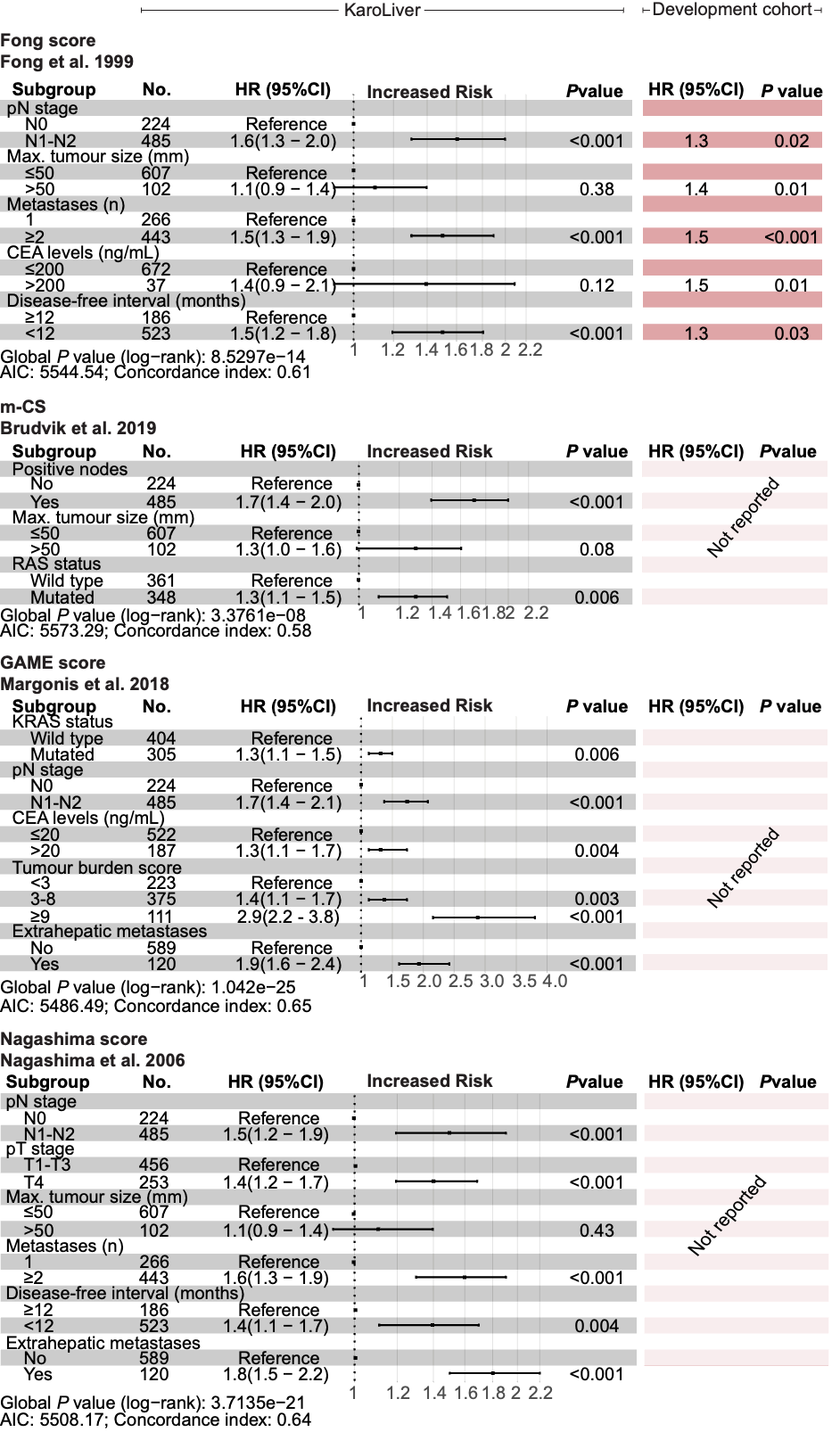

Total number of events in KaroLiver: 476. Hazard ratios (HR) and *P* values are derived from multivariable Cox proportional hazard regression. Results from development cohorts (i.e. original publications) are provided where available. AIC, Akaike information criterion; CEA carcinoembryonic antigen.

**Fig. S6 Multivariable Analyses of Konopke, Helsinki, and Nordlinger Score Risk Factors in KaroLiver (Recurrence-Free Survival)**

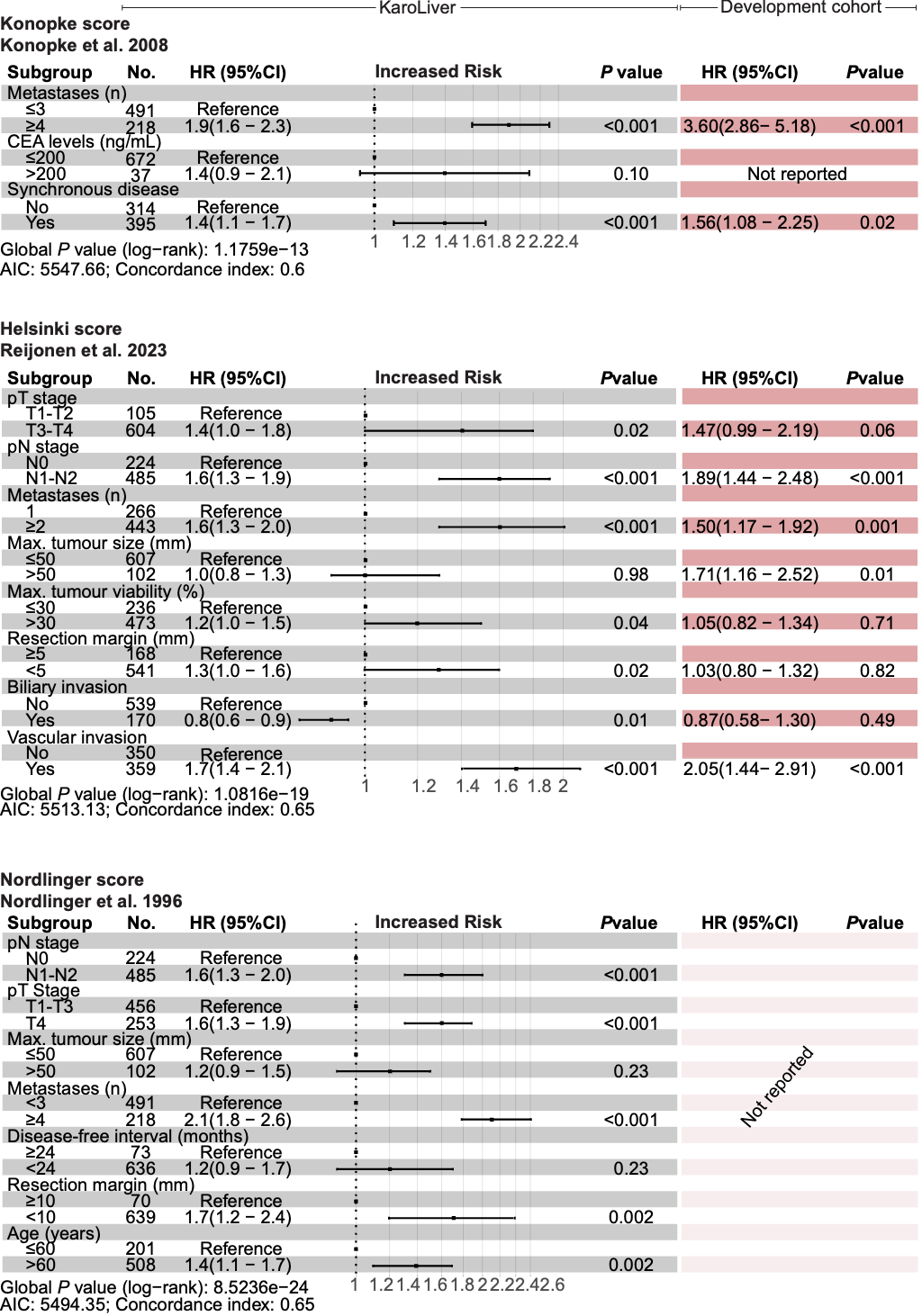

Total number of events in KaroLiver: 476. Hazard ratios (HR) and *P* values are derived from multivariable Cox proportional hazard regression. Results from development cohorts (i.e. original publications) are provided where available. AIC, Akaike information criterion; CEA carcinoembryonic antigen.

**Fig. S7 Multivariable Analyses of Fong, m-CS, GAME, and Nagashima Score Risk Factors in KaroLiver (Overall Survival)**

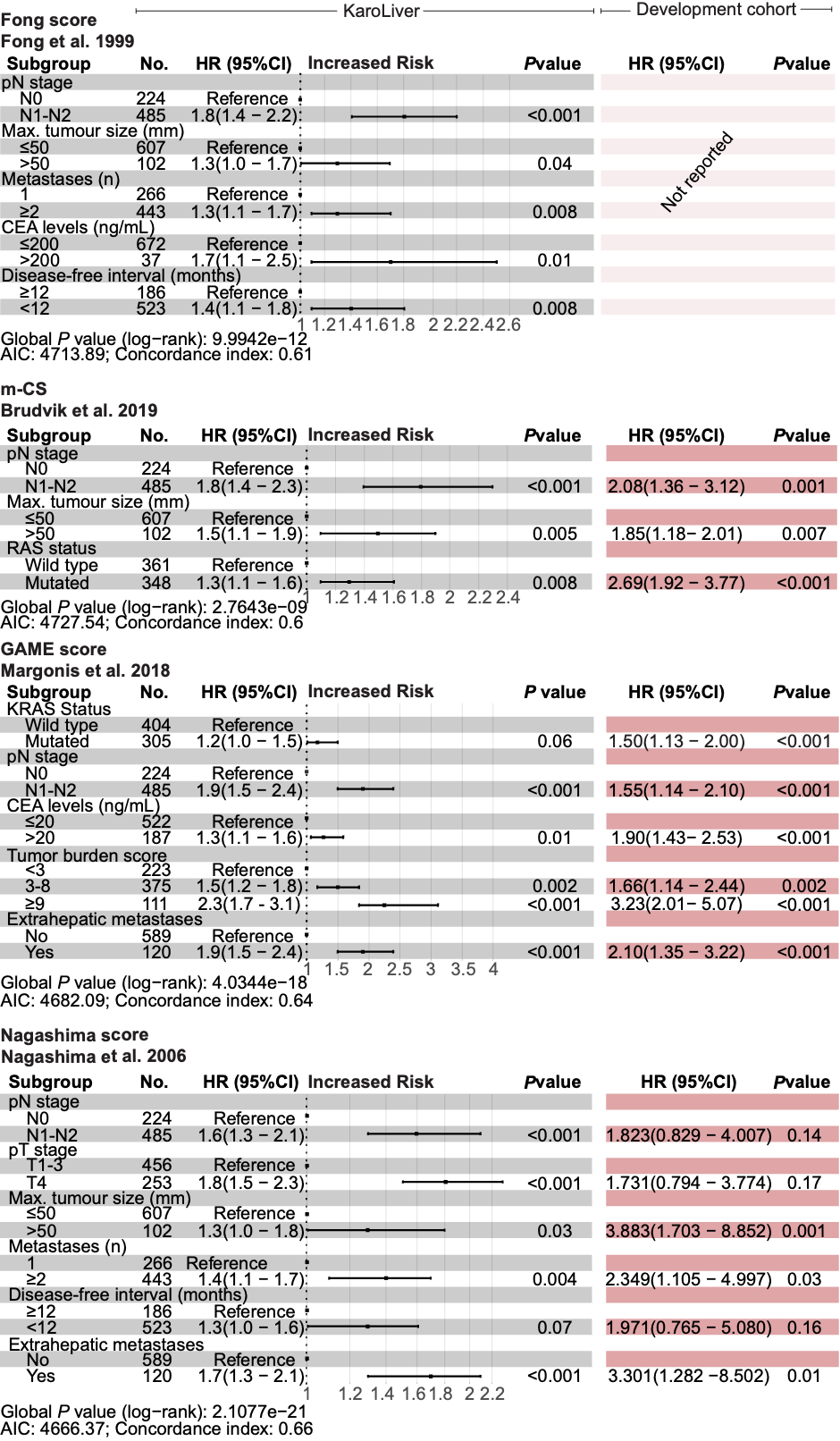

Total number of events in KaroLiver: 395. Hazard ratios (HR) and *P* values are derived from multivariable Cox proportional hazard regression. Results from development cohorts (i.e. original publications) are provided where available. AIC, Akaike information criterion; CEA, carcinoembryonic antigen.

**Fig. S8. Multivariable Analyses of Konopke, Helsinki, and Nordlinger Score Risk Factors in KaroLiver (Overall Survival)**

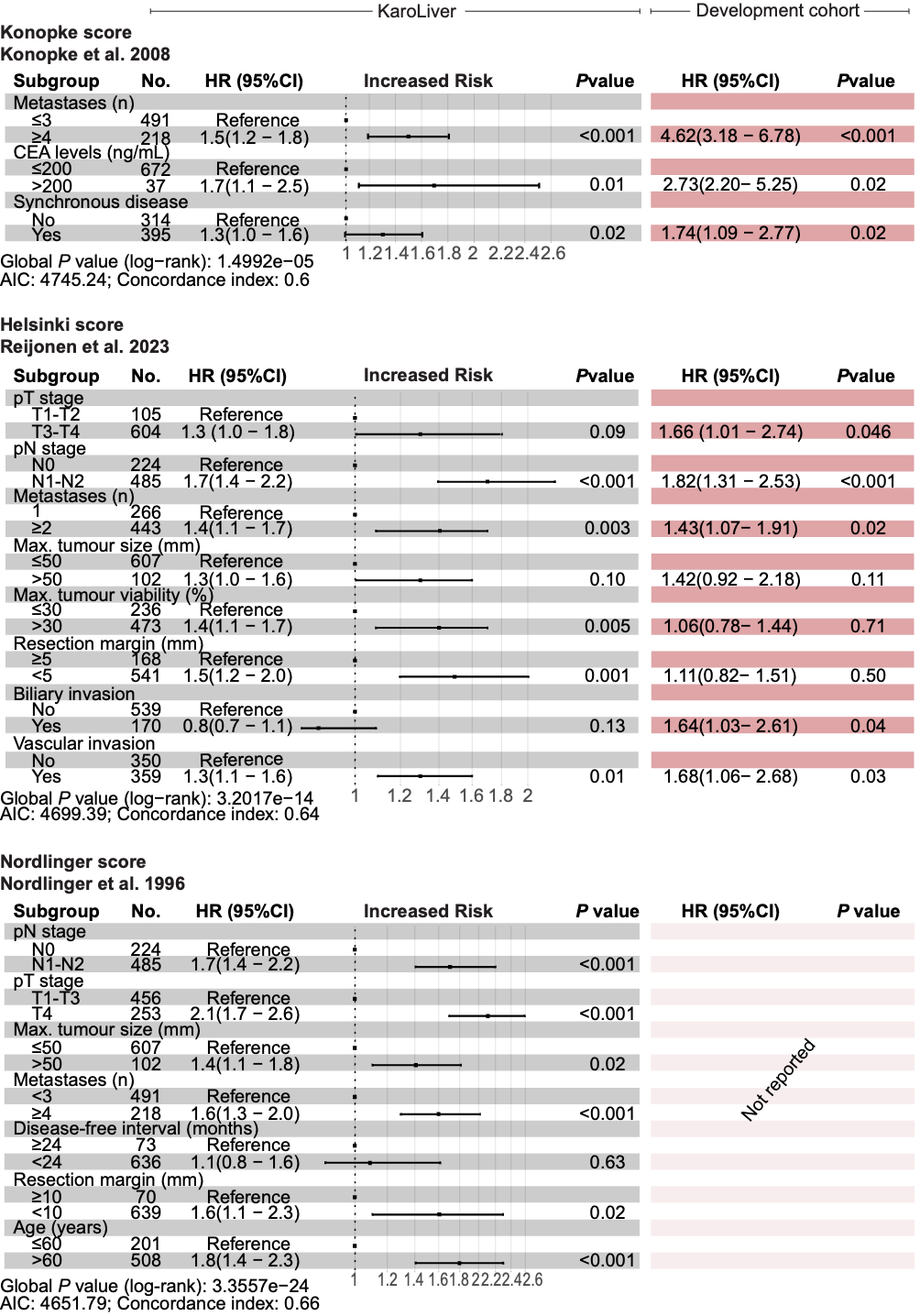

Total number of events in KaroLiver: 395. Hazard ratios (HR) and *P* values derived from development cohorts are from multivariable Cox proportional hazard regression. Results from development cohorts (i.e. original publications) are provided where available. AIC, Akaike information criterion; CEA, carcinoembryonic antigen.

**Fig. S9 Calibration of Recalibrated Clinical Risk Score Models in KaroLiver**

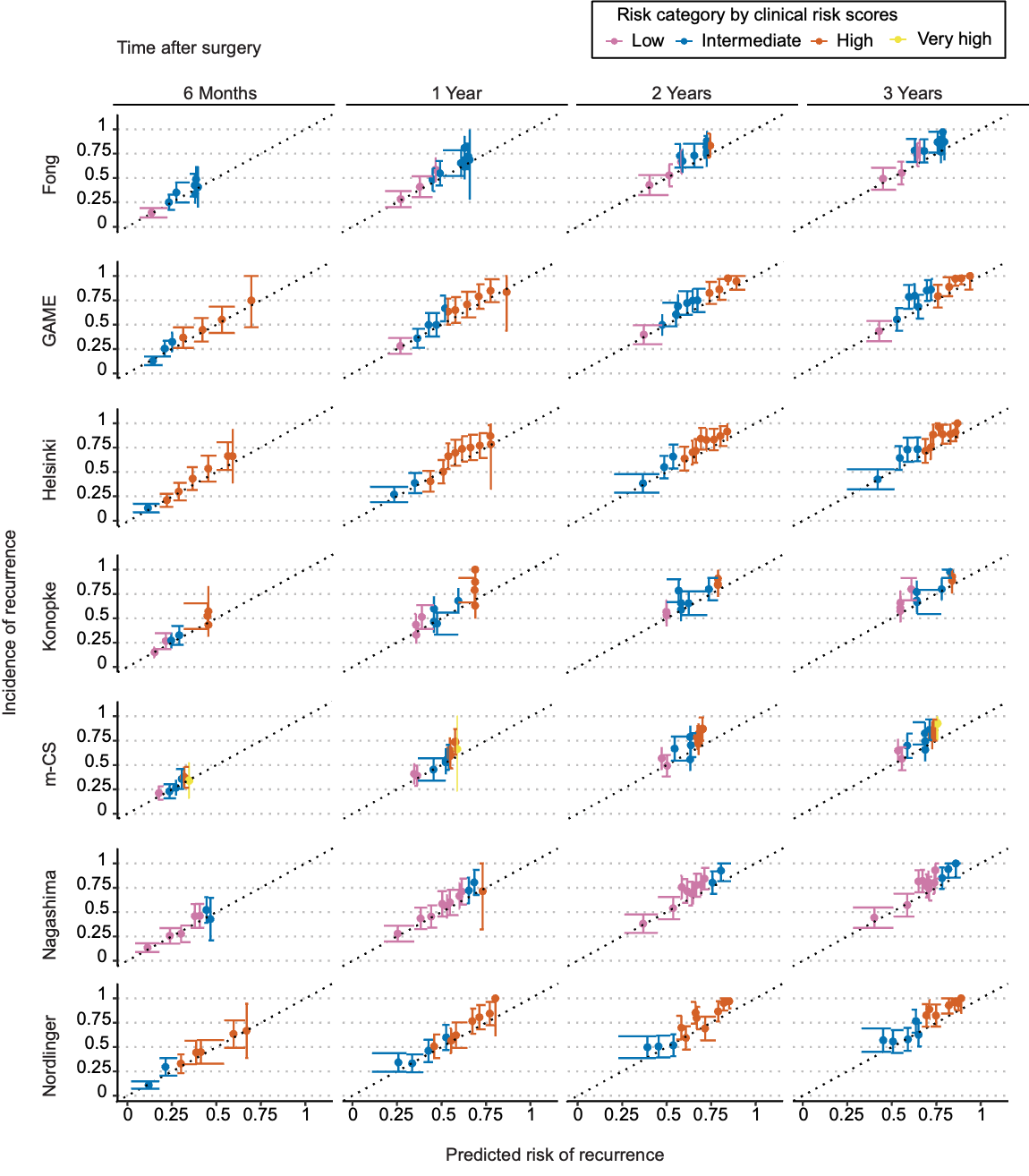

Each dot represents the mean predicted risk of recurrence after colorectal cancer liver metastasis resection compared with the observed incidence (95% CI, error bars). Diagonal dotted lines represent perfect alignment between the observed and predicted risks. GAME, genetic and morphological evaluation; m-CS, modified clinical risk score*.*

**Fig. S10 Calibration of Recalibrated Clinical Risk Score Models after Bootstrap Internal Validation in KaroLiver**

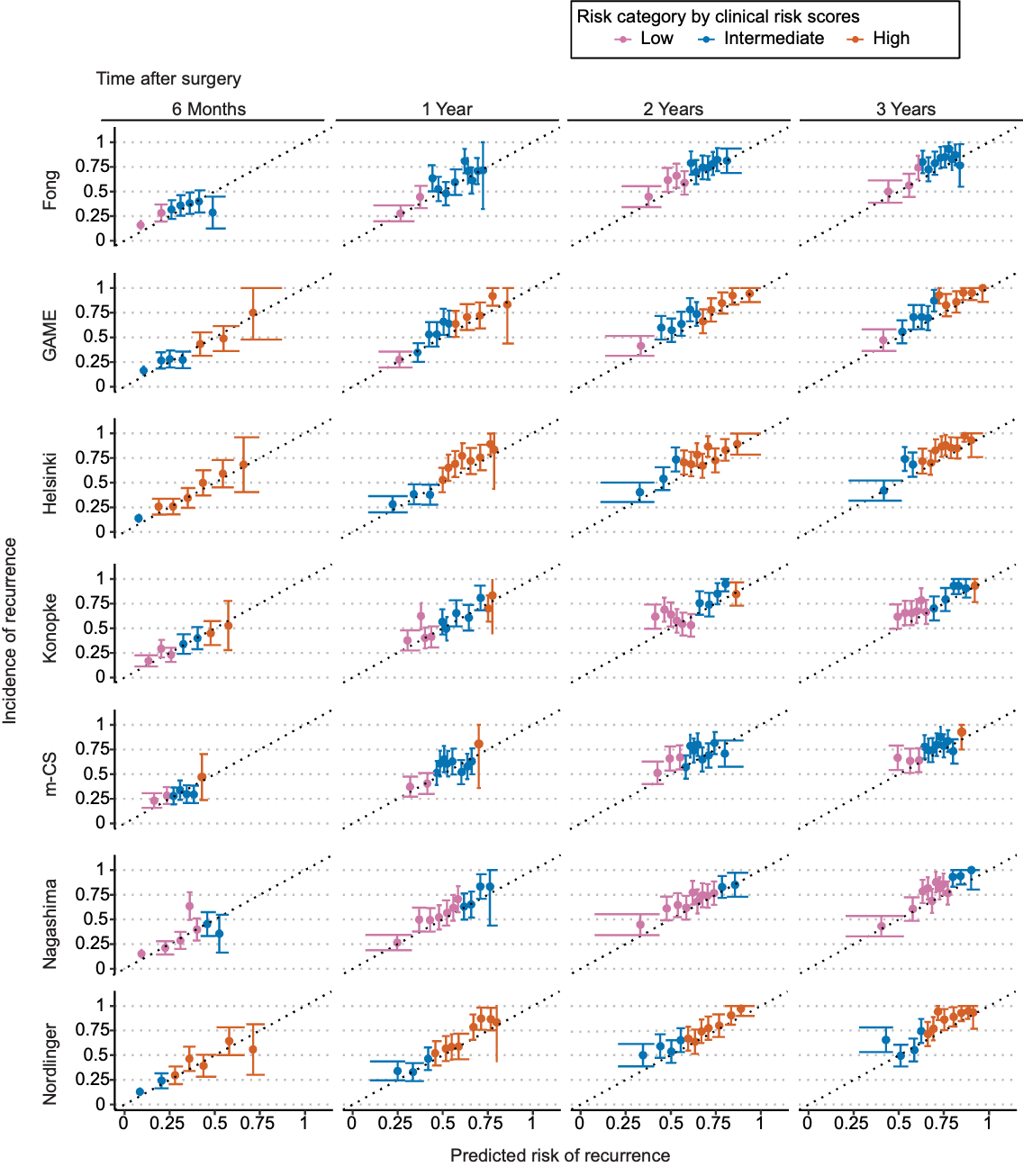

Each dot represents the mean predicted risk of recurrence after colorectal cancer liver metastasis resection compared with the observed incidence (95% CI, error bars). Diagonal dotted lines represent perfect alignment between the observed and predicted risks. GAME, genetic and morphological evaluation; m-CS, modified clinical risk score.

**Fig. S11 Decision Curve Analysis for Recalibrated Postoperative Scoring Models with and without Postoperative Factors in KaroLiver**

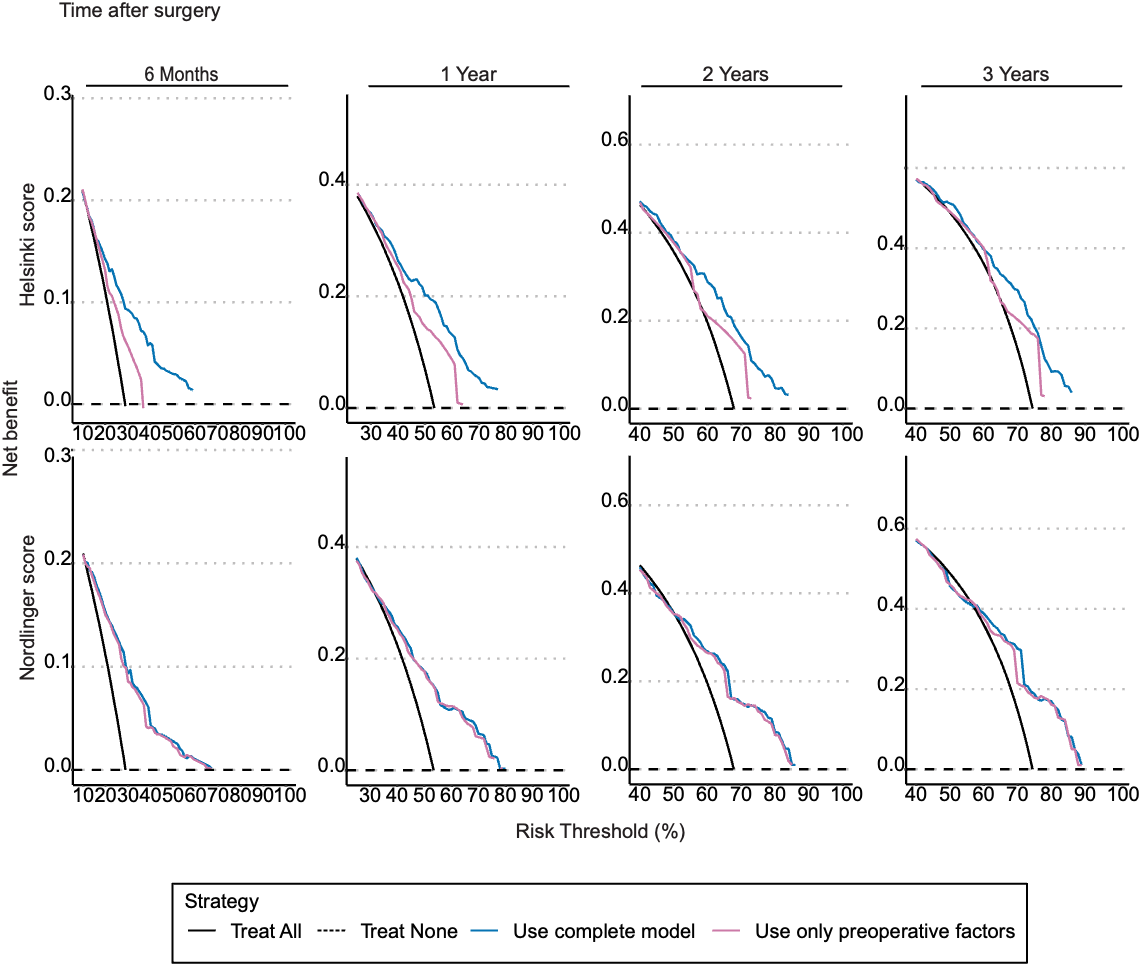

A higher net benefit than the “treat all” scenario indicates a better balance between true and false positives with the scoring model. The x-axis represents the threshold probability for treatment. CRLM, colorectal cancer liver metastasis. **References**
